# Large-scale association study identifies lung cancer susceptibility copy number variants and their potential functional role in genetic instability

**DOI:** 10.64898/2026.05.11.26352741

**Authors:** Feifei Xiao, Fei Qin, Xizhi Luo, Shannon E Slewitzke, Gail F Fernandes, Mattias Johansson, Xiangjun Xiao, David Zaridze, Stig Egil Bojesen, Sanjay Shete, Demetrios Albanes, Melinda C. Aldrich, Adonina Tardon, Guillermo Fernandez-Tardon, Loïc Le Marchand, Gad Rennert, Heike Bickeböller, H-Erich Wichmann, Angela Risch, Thomas Muley, Albert Rosenberger, John K. Field, Michael Davies, Penella Woll, Lambertus A. Kiemeney, Aage Haugen, Shanbeh Zienolddiny, Stephen Lam, Mikael Johansson, Kjell Grankvist, Matthew B. Schabath, Angeline Andrew, Philip Lazarus, Susanne M. Arnold, Dakai Zhu, Hermann Brenner, Marian L. Neuhouser, Rayjean J. Hung, David C. Christiani, James McKay, Guoshuai Cai, Jun Xia, Christopher I. Amos

**Author notes:** Corresponding author information: Christopher I. Amos, Ph.D., Professor, University of New Mexico Health Sciences, Associate Director for Population and Data Science, UNM Comprehensive Cancer Center Head, Data Science Initiatives.

## Abstract

**Background:** Genome-wide association studies (GWAS) have identified numerous lung cancer susceptibility loci based on single nucleotide polymorphisms (SNPs), yet a substantial proportion of heritability remains unexplained. We therefore evaluated germline copy number variants (CNVs) as an underexplored source of genetic susceptibility and potential contributors to genomic instability in lung cancer.

**Methods:** We conducted a genome-wide analysis of germline CNVs using 19,342 cases and 15,917 controls from the Transdisciplinary Research in Cancer of the Lung (TRICL) consortium, with replication in two independent cohorts. High-confidence CNVs were identified by integrating two CNV callers including PennCNV and modSaRa2. Association analyses were performed using both gene-based and CNV region–based approaches. Polygenic risk scores (PRS) were constructed from top loci, and functional validation was conducted using siRNA-mediated knockdown in lung fibroblast cells.

**Results:** We identified CNVs in four genomic regions (1p36.22, 2q31.2, 6p21.32, and 19q13.32) significantly associated with lung cancer risk. Two loci (1p36.22 and 2q31.2) were consistently supported across both analytical strategies. A CNV-based PRS constructed from key genes (CLCN6, NFE2L2, OPA3, and PSMB8) was significantly associated with lung cancer risk and replicated across independent datasets. Functional assays demonstrated that knockdown of NFE2L2 and OPA3 increased endogenous DNA damage, supporting a role in genomic stability.

**Conclusions:** Germline CNVs contribute to lung cancer susceptibility and may influence carcinogenesis through mechanisms related to genomic instability.

**Impact:** These findings expand the genetic architecture of lung cancer and highlight CNVs as potential biomarkers for improving risk stratification and informing precision prevention strategies.

## Introduction

Lung cancer (LC) remains the leading cause of cancer-related mortality worldwide, accounting for substantial global morbidity and mortality (1). Despite advances in early detection and targeted therapies, prognosis remains poor, largely because most cases are diagnosed at advanced stages. Although tobacco smoking is the primary risk factor, up to 25% of lung cancer cases occur in never smokers, underscoring the contribution of additional etiologic factors, including inherited genetic susceptibility (2). Twin studies estimate the heritability of LC to be approximately 18% (3), yet a substantial proportion of this genetic risk remains unexplained.

Genome-wide association studies (GWAS) based on single nucleotide polymorphisms (SNPs) have identified over 50 susceptibility loci for lung cancer, including well-established regions such as 5p15.33, 6p21–6p22, and 15q25.1 (4-13). While these discoveries have advanced our understanding of lung cancer genetics, they explain only a fraction of heritable risk, suggesting that additional forms of genomic variation contribute to disease susceptibility. Structural variants, particularly copy number variants (CNVs), represent a major but underexplored source of genetic variation. CNVs defined as deletions or duplications of genomic segments, can influence gene dosage, disrupt regulatory elements, and alter chromatin architecture, thereby exerting substantial functional effects (14,15). Consistent with their biological importance, copy number alterations affect a large proportion of cancer-related genes (16).

Accumulating evidence implicates CNVs in the etiology of complex diseases, including autoimmune (17,18), neurological (19,20), and oncologic conditions (21-23). However, the role of germline CNVs in lung cancer susceptibility remains poorly characterized, with existing studies limited to small-scale analyses or focused on somatic alterations in tumors rather than inherited variation [21,22]. This represents a critical gap in understanding the genetic architecture of lung cancer. Importantly, CNVs may contribute not only to disease susceptibility but also to genomic instability, a hallmark of cancer that underlies tumor initiation and progression (24,25).

Several GWAS-identified susceptibility regions, such as 5p15.33, 6p21.32, and 15q25.1, have been consistently replicated across populations (11,13,26,27). In addition, alterations in genes such as *NFE2L2* (2q31.2), including copy number gains and activating mutations, have been implicated in lung tumorigenesis through dysregulation of oxidative stress pathways (28-30). However, these findings primarily derive from tumor-based analyses or SNP-level studies, rather than direct evaluation of germline CNVs and their contribution to inherited risk.

To address this gap, we leveraged data from the Transdisciplinary Research in Cancer of the Lung (TRICL) consortium, one of the largest available lung cancer genetic resources, to perform a comprehensive genome-wide analysis of germline CNVs. We integrated complementary CNV-calling algorithms to enhance detection reliability and applied both gene-based and region-based association approaches. Findings were evaluated in independent validation datasets, and cumulative genetic effects were assessed using CNV-based polygenic risk scores. In addition, functional assays in a lung fibroblast cell model were conducted to investigate the potential role of candidate genes in promoting endogenous DNA damage.

Our objectives were to (1) identify germline CNVs associated with lung cancer susceptibility, and (2) investigate their potential functional roles in promoting genomic instability. By systematically interrogating CNVs at a genome-wide scale, this study provides new insights into the genetic architecture of lung cancer and highlights previously underrecognized mechanisms linking inherited variants to carcinogenesis.

## Methods

### Datasets

This study was conducted using the OncoArray datasets from the Transdisciplinary Research in Cancer of the Lung (TRICL) as part of the International Lung Cancer Consortium (ILCCO). Details of the participating studies and genotyping protocols have been previously described in the OncoArray GWAS study (13). Briefly, the primary discovery dataset comprised 19,342 lung cancer (LC) cases and 15,917 controls. DNA sampled from whole blood were collected and 533,631 single nucleotide polymorphisms (SNPs) were genotyped by the OncoArray, which was a special array configured for understanding genetic factors influencing common cancers (31). Demographic and lifestyle data including race, sex, age, histological subtypes, and cigarettes smoking status were systematically collected.

For validation analyses, two additional lung cancer cohorts were employed including: (1) a German OncoArray cohort comprising 1,851 individuals, and (2) an Affymetrix microarray study involving 6,035 participants (32). The German OncoArray samples were drawn from a large lung cancer study of 3,354 individuals that was separately genotyped from the rest of the OncoArray subjects by researchers in Heidelberg along with additional samples that were genotyped after the release of the OncoArray project, which included subsets labeled “GERMANY-1344”, “GERMANY-1680”, and “NEWSAMPLE-600”. After exclusion of overlapping samples with the TRICL-OncoArray study and phenotype-based filtering, 1,851 remained for validation in this study. The Affymetrix dataset was derived from an initial cohort of 12,651 individuals. After removing samples overlapping the OncoArray cohort, 6,035 independent samples remained for our validation study. Samples from these two validation datasets were all of European ancestry.

### Sample and Genotyping Quality Control (QC)

Raw intensity data (.idat files) were processed using Illumina GenomeStudio (v2.0.3) to generate Log R Ratio (LRR) and B allele frequency (BAF) values. Pre-calling quality control (QC) filters were applied including exclusion of samples with (1) unexpected duplicates or close relatedness (i.e., identity by descent > 0.2); (2) low genotyping call rates (i.e., call rate < 0.95); (3) excessive LRR variation (i.e., LRR standard deviation [LRR_SD] > 0.35 on chromosome 1) (33); and (4) B allele frequency drift > 0.01; (5) discordant sex. PLINK 1.9 (34) was used to exclude duplicates or relatives. Identity by descent was calculated between pairs of samples to detect unexpected duplicates 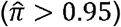 or relatedness 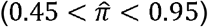. Complete genotype data for X chromosomes was used to verify reported sex by using PLINK sex inference.

Given that structural variants such as duplications or deletions may introduce bias in genotyping calls or departure from Hardy–Weinberg equilibrium (HWE), no SNP-level quality control procedure was applied (35). After sample and genotyping QC, there were 7,544 lung adenocarcinoma (LUAD) patients, 4,468 squamous carcinoma (LUSC), 1,719 small cell lung cancer (SCLC) and the others were unknown subtypes (Supplementary Table S1).

### CNV Calling and Post-calling QC

CNV calling was jointly performed using PennCNV (36) and modSaRa2 (37). Method-specific post-calling QCs were applied based on CNV length, copy number state, and confidence metrics (Supplementary Figure S1). CNVs were excluded if they (1) overlapped with centromeres; (2) contained fewer than 10 SNPs; or (3) exceeded 300kb (for calls generated by PennCNV), or (4) exhibited confidence scores below 10 (for calls generated by PennCNV). CNVs were then annotated using human GRCh37/hg19 reference genome build. Each CNV was labeled with the associated gene, chromosomal location, position in marker index, length and copy number state (duplications, diploids/normal or deletions). Regarding gene inclusion, no genes were excluded due to lack of SNP coverage for CNV determination. CNV sets from these two algorithms were merged to optimize CNV calling validity. Visual inspection of randomly selected CNVs from both callers via LRR and BAF visualization plots validated the quality of the merged set. Comparative analyses demonstrated that modSaRa2 showed superior sensitivity for long CNVs (>200 kb) (Supplementary Table S2), and these calls were preferentially integrated to the PennCNV profiling results.

### Genome-wide CNV and Lung Cancer Risk Association

We employed two approaches: (1) gene-based association testing as the primary analysis using gene coding regions as the testing units, and (2) CNV-region based association testing using CNVruler (38) to define the CNV regions (CNVRs). For gene-based association analyses, CNVs were mapped to gene coding regions as defined by our annotation, and their effects were assessed. CNVs located upstream of a gene including those potentially affecting promoter or regulatory regions, were not assigned to that gene and were excluded in the analyses. We evaluated the associations between deletions (i.e., copy number loss), duplications (i.e., copy number gain), or presence of CNVs (deletions or duplications) and lung cancer risk.

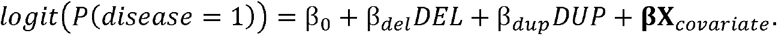

Stratification analyses were conducted by histological subtypes (in the whole population [overall], LUAD, LUSC and SCLC) and by the genetically inferred ancestry (overall and in the European ancestry group, which was the largest subpopulation). Demographic information including age, sex and smoking status, study site and four ancestry related principal components [PC] were adjusted in all regression models. The definition of genetically inferred ancestry is described in the original GWAS study of OncoArray (31). Individuals were classified into European (defined as >80% European ancestry), Asian (>40% Asian ancestry), African (>20% African ancestry) and other.

For the CNVR-based association, CNV regions were defined using CNVRuler (38). CNVRuler is a robust method which aligned consensus CNV signals of multiple individuals into regions, where the boundaries of CNVs were determined by the observed data. With the high-confidence CNV calls, CNVRuler was used to merge overlapping CNV calls across samples (at least 1 base-pair overlapping) and it was also simultaneously used to trim the low-frequency regions (<10%). Long CNVRs exceeding 500kb in length were excluded. CNVR-based strategy allowed the investigation of large genomic regions that covered multiple genes. The association modeling was mirroring the gene-based association framework. For all analyses, genome-wide significance was set at P < 5.6 × 10^−6^ after Bonferroni correction (as computed in the CNVR approach). Thse same p-value threshold was used in the gene-based approach (relaxed compared with the original 2.3 × 10^−6^ after Bonferroni correction), overall and the subtype-specific analyses.

### Validation and Sensitivity Analyses

Two validation datasets were used including the German OncoArray (n=1,851) and an Affymetrix microarray dataset (n=6,035). To assess the robustness of the lung cancer susceptibility genes, sensitivity analyses were also performed in the Asian samples from the TRICL-OncoArray dataset, Chinese Han specifically (N=2,826). Data normalization, pre- and post-CNV calling QC steps, CNV calling methods, and association pipelines remained the same as those in the TRICL discovery dataset. The generated CNVs were annotated using the same reference genome build as the discovery data. Considering the relatively small sample sizes in the validation and sub-population datasets, Fisher’s exact test was applied to test the association when the CNV counts in groups were fewer than ten.

### Polygenic Risk Scores Analysis

The Polygenic risk score (PRS) has proven to be an efficient approach to study complex diseases by integrating a curated set of disease-associated variants, often explaining a greater proportion of disease risk than a single variant. PRS were constructed from significant CNVs in the TRICL GWAS gene-based association analysis. Note that PRS defined in this study used the CNVs instead of single nucleotide variants as in standard PRS approach. Three PRS calculation methods were used including: (1) weighted score using reported risk variant effect sizes (*PRS*_*β*_); (2) unweighted sum of risk variants (*PRS*_*uw*_); and (3) inverse variant weighted scores incorporating the standard error of the effect sizes (*PRS*_*IV*_).

First, we used the standard PRS weights, corresponding to the reported CNV effect sizes in which the log odds ratio (*β*) was obtained from the TRICL-GWAS gene-based association study:

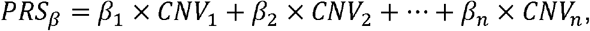

where *CNV*_*i*_ (*i* = 1, … *n*) is an indicator function about the existence of the risk variant (e.g., deletion). We compared this to an unweighted score computed from the sum of the risk variants, which assigns an equal weight of 1 to each CNV, irrespective of effect sizes:

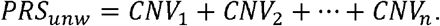

Lastly, to account for uncertainty in estimating risk variant effect sizes and down-weight imprecisely estimated effect sizes, inverse variance (IV) weights were applied in the PRS calculation that incorporated the standard error (SE) of the estimated log odds ratio (*β*) from the TRICL-GWAS study:

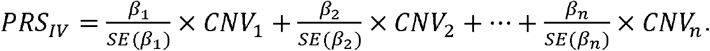

Details of these PRS approaches in constructing SNP based PRS was descried in Kachuri *et al*., 2020 (39). Each PRS was then standardized based on its mean and standard deviation and then evaluated in a logistic regression to assess the effect of the computed PRS in predisposing to LC overall risk, adjusting for the same covariates as in the primary TRICL-OncoArray genome-wide CNV association study. We also computed PRS using an expanded set of the top twenty variants. For pruning of the CNV sets, independent CNVs were prioritized by selecting one representative CNV per genomic cytoband based on the smallest p-value to avoid redundancy from overlapping CNV events.

Using the same genetic variants set as in the discovery dataset, PRS validation analyses were conducted in the two validation datasets, with PRS computed from effect sizes derived from each of these cohorts, respectively. We acknowledge that effect sizes derived from each dataset may be subject to the “winner’s curse”, potentially leading to biased estimates. However, the use of independent loci and subsequent validation helps mitigate this concern.

### Pathway and Network Analysis

To elucidate potential biological pathways linked to lung cancer susceptibility, gene set enrichment analysis (GSEA) (40) was performed on the top 200 CNV-associated genes detected in the gene-based and CNVR-based association analyses. Enrichment was tested against KEGG (41) and MsigDB hallmark pathways (42). A false discovery rate (FDR) threshold of < 0.05 was applied to declare statistical significance.

### Cell Line and Reagents

The MRC5-SV40 human lung fibroblast cell line (male, SV40-immortalized, source: Dr. Stephen P. Jackson Lab via Dr. Kyle Miller) was cultured in DMEM high glucose medium (Gibco, #11965118) with 10% fetal bovine serum (Gibco, #10438034), 2mM L-glutamine, and antibiotics (100ug/ml streptomycin and 100 ug/ml penicillin, Gibco, #10378016). It underwent short tandem repeats (STR) analysis authentication (ATCC, July 2018), regular mycoplasma testing (ABM, G238). The cells used in this study were directly derived from the cell population that underwent STR analysis.

### DNA Damage Assays

DNA damage was quantified using γH2AX-based flow cytometry 72 hours post-transfection with siRNA smartpools or individual siRNAs. Staining involved γH2AX primary antibody (Sigma, Catalog #05-636) and Alexa Fluor 647-conjugated goat anti-mouse secondary antibody (Thermo Fisher, Catalog #A21236). A BD LSRFortessa flow cytometer analyzed stained cells, with FlowJo software processing the FCS files. Post 72-hour siRNA transfection, median fluorescence intensity of the cells and the percentage of γH2AX positive cells were quantified, compared to non-targeting siRNA controls. Non-targeting pool siRNA (D-001810-10), SMARTpool siRNAs each containing four targeting sequences of *CLCN6, NFE2L2, OPA3, PPM1N, PSMB8, RTN2, VASP*, and sets of 4 siRNAs targeting *OPA3 and NFE2L2* were purchased from Dharmacon. The target sequences for *OPA3 and NFE2L2* are as follows: #1 OPA3 (GCGCGUUCCCUAUGGCGAA), #2 OPA3 (UGGCGAAGCUGCUAUACUU), #3 OPA3 (GCAUCCGGCAGGUCAGCAA), #4 OPA3 (CCGCUUGCCAACCGUAUUA); #1 NFE2L2 (UAAAGUGGCUGCUCAGAAU), #2 NFE2L2 (GAGUUACAGUGUCUUAAUA), #3 NFE2L2 (UGGAGUAAGUCGAGAAGUA), #4 NFE2L2 (CACCUUAUAUCUCGAAGUU). siRNA transfections were carried out with lipofectamine RNAiMax Transfection Reagent (#13778075, Invitrogen), following the manufacturer’s recommendations. SMARTpool ON-TARGETplus siRNA was designed and modified for greater specificity and reduce off-targets up to 90% utilizing a dual-strand modification.

### Real-time quantitative reverse transcription PCR (RT-qPCR), siRNA targeting sequences,and primers

Total RNA was extracted from cells 72□h post siRNA transfection using the RNeasy mini kit (Qiagen #74106). 300□ng of total RNA from each sample was used to synthesize cDNA by the Superscript III first-strand synthesis system (Invitrogen, #18080051). The qPCR reactions were performed using iTaq Universal SYBR Green Supermix (BioRad #172-5121) on a QuantStudio 3 Real-Time PCR System (Applied Biosystems). For each gene, three replicates were analyzed, and the average threshold cycle (Ct) was calculated. The relative expression levels were calculated with the 2–ΔΔCt method. Knockdown efficiency was quantified by RT-qPCR and shown below. In the assessment of siRNA knockdown efficiencies using SMARTpool, all tested genes exhibited notable silencing efficacies. *OPA3* demonstrated a knockdown efficiency of 93.51%, *VASP* at 89.72%, *NFE2L2* at 82.31%, *PSMB8* at 91.36%, *CLCN6* at 97.66%, *PPM1N* at 61.17%, and *RTN2* at 85.35%. Collectively, these data suggest an overall high level of knockdown efficiency across the tested gene targets, affirming the efficacy of SMARTpool-mediated siRNA in gene silencing applications. Each of the four individual siRNAs targeting *NFE2L2* and *OPA3* was tested for efficiency. The knockdown efficiency was as follows: NFE2L2 #1: 77.43%, NFE2L2 #2: 77.11%, NFE2L2 #3: 65.81%, NFE2L2 #4: 53.44%, OPA3 #1: 78.96%, OPA3 #2: 73.97%, OPA3 #3: 56.99%, and OPA3 #4: 66.54%. Altogether, NFE2L2 #1, #2 and OPA3 #1, #2 were selected for further DNA damage validation by individual siRNAs based on their high knockdown efficiencies. Primers used included GAPDH (housekeeping gene) forward: CAA TGA CCC CTT CAT TGA CC; GAPDH reverse: GAT CTC GCT CCT GGA AGA TG; CLCN6 forward: TTC ACC CAA CTC AAG TTC GGA; CLCN6 reverse: ACC GGC TCA ATG AGA ACA AGG; NFE2L2 forward: CCT ATG GCG AAG CTG CTA TAC; NFE2L2 reverse: GGC GGC CTC CTT AAT ACG G; PPM1N forward: CGA GCG TTG GGC GAC TTT A; PPM1N reverse: CAG GAG CAT GAA CTC GTC CT; RTN2 forward: GAC CTG CTG TAC TGG AAG GAC; RTN2 reverse: ACG GAC ACG ATG CTA AAG TGC; VASP forward: ATG GCA ACA AGC GAT GGC T; VASP reverse: CGA TGG CAC AGT TGA TGA CCA; OPA3 forward: CCT ATG GCG AAG CTG CTA TAC; OPA3 reverse: GGC GGC CTC CTT AAT ACG G; PSMB8 forward: CAC GCT CGC CTT CAA GTT C; PSMB8 reverse: AGG CAC TAA TGT AGG ACC CAG.

## Results

### Genome-wide high confidence CNV calling

We undertook a genome-wide CNV calling using OncoArray data from the TRICL consortium, which genotyped 533,631 SNPs (Methods and Supplementary Table S1), primarily from individuals of European ancestry (87.5%). After genotyping quality control, a total of 18,455 lung cancer cases and 15,065 controls remained. Accurate *in silico* identification of relatively short germline CNVs remains a methodological challenge. To enhance the validity, we systematically integrated two CNV detection algorithms, PennCNV (36) and modSaRa2 (35), to derive high-confidence CNV calls. Empirically, modSaRa2 demonstrated high sensitivity, whereas PennCNV showed high consistency and reliability. A summary of the CNV quality visualization for each CNV caller (Supplementary Table S2) suggested that the two CNV sets be merged.

Consequently, 1,243,335 CNV calls were generated across 21,402 known autosomal genes (Supplementary Figure S1). No difference was observed between cases and controls in the number of CNVs per individual, CNV length distribution, or quality metrics including Log R Ratio (LRR) standard deviation or B allele frequency (BAF) drift (Supplementary Table S3; Supplementary Figure S2). Even without using standard SNP/probe-level QC filters, representative LRR and BAF plots at the top loci (Supplementary Figure S3) demonstrate consistent intensity patterns between carriers and non-carriers, supporting the validity of the identified CNVs.

### Genome-wide CNV analysis identifies lung cancer susceptibility loci with TRICL dataset

Two strategies were used in genome-wide CNV association including gene based and CNVR based association (Methods), from which overlapping significant results were considered as high-confidence findings. Manhattan plots by histology and population ancestry for the two association analyses are displayed in Figures 1-2. No evidence of genomic inflation was observed in the overall analysis or subtype-specific analysis, indicating minimal confounding from hidden population structure (Supplementary Figure S4). A full list of overall, subtype-specific, and stratification association results is shown in Supplementary Tables S4-5.

**Fig. 1.**
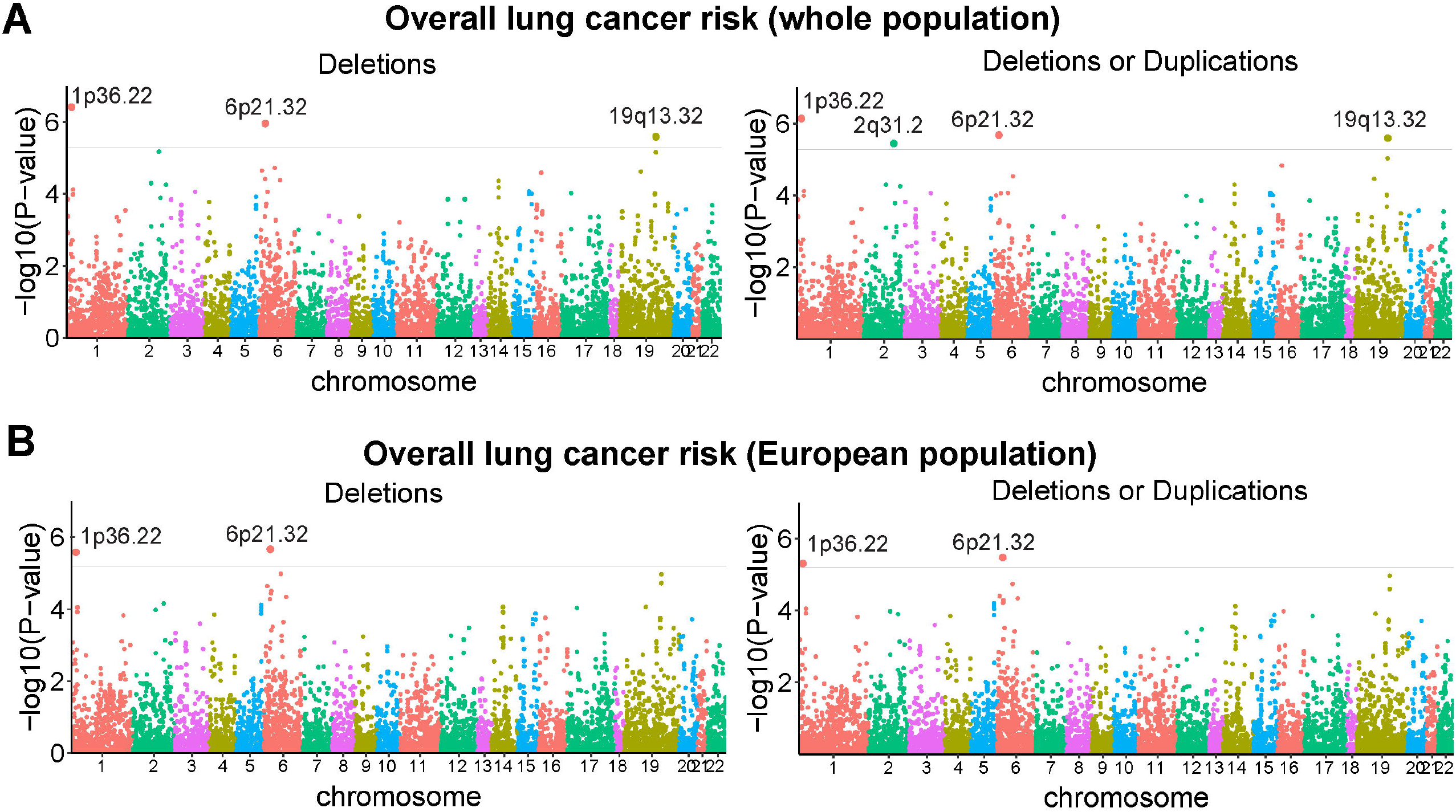
Manhattan plots for the gene-based association analysis of lung cancer and stratification analysis by histological subtype and ethnicity. (A) Overall lung cancer risk in the whole population: 18,455 cases and 15,065 controls. (B) Overall lung cancer risk in the European ancestry population: 16,556 cases and 12,768 controls. Each significant locus is annotated by its cytoband location. The x axis represents chromosomal coordinates, and y axis represents corresponding P-values (-log_10_ scale). The red horizontal line is the statistical significance threshold P-value= 5.6× 10^−6^.

**Fig. 2.**
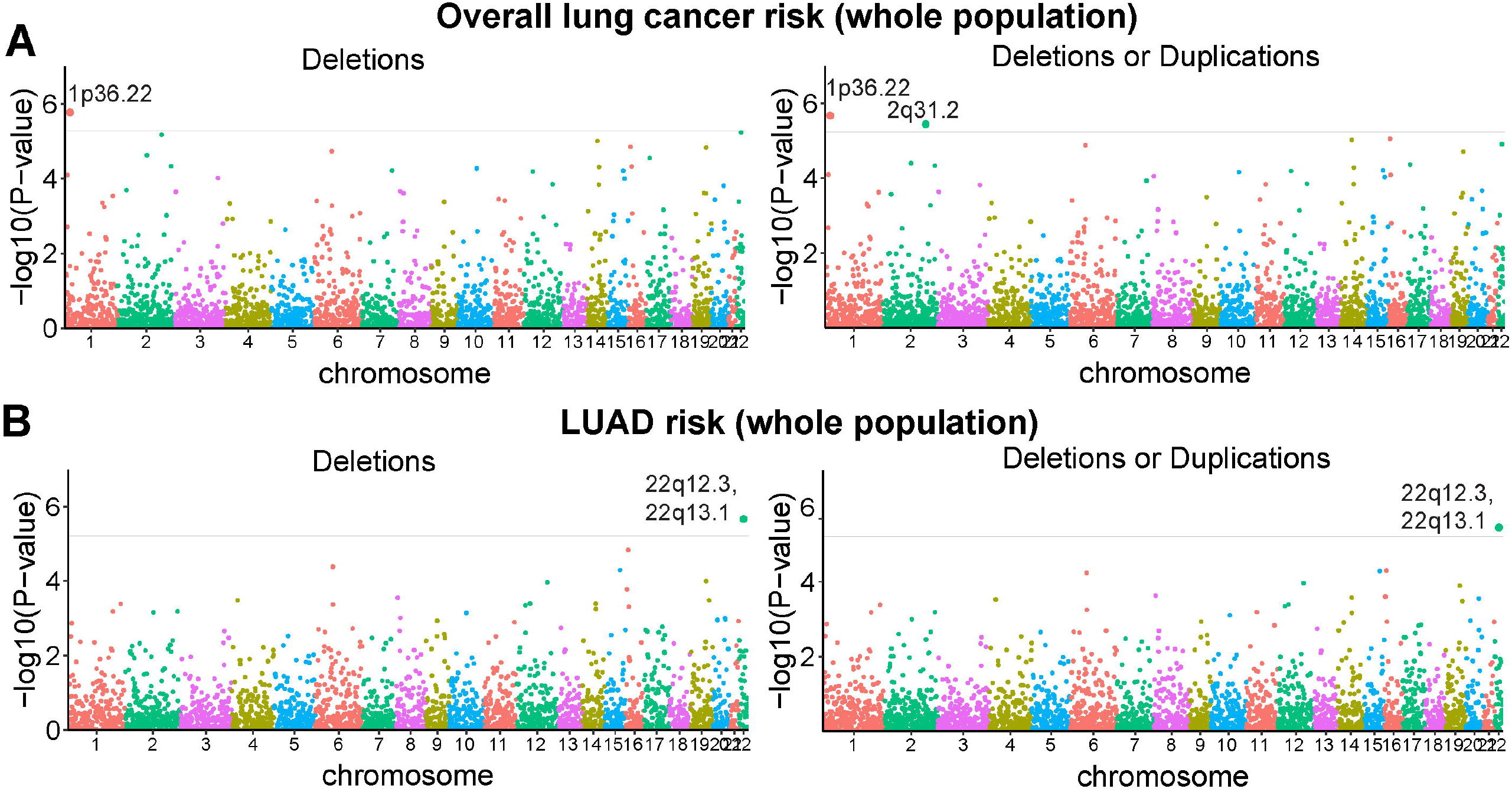
Manhattan plots for the CNV region (CNVR)-based genome-wide association analysis of lung cancer risk and stratification analysis by histological subtype and ethnicity. (A) Overall lung cancer risk in the whole population: 18,455 cases and 15,065 controls. (B) Adenocarcinoma risk in the whole population: 7,544 cases and 15,065 controls. Each significant locus is annotated by its cytoband location. The x axis represents chromosomal coordinates, and y axis represents corresponding P-values (-log_10_ scale). The red horizontal line is the statistical significance threshold P-value= 5.6 × 10^−6^.

For gene-based association analysis, all significant findings were reported for LC overall and by histological subtypes (Table 1, Figure 1). Deletions in four genes located at four genomic regions were suggestively associated, including *CLCN6* at 1p36.22 (Deletion OR=1.82, 95% CI=1.45-2.30), *NFE2L2* at 2q31.2 (Deletion OR=2.01, 95% CI=1.48-2.72), *OPA3* at 19q13.32 (Deletion OR=2.63, 95% CI=1.76-3.93) and *PSMB8* at 6p21.32 (Deletion OR=1.94, 95% CI=1.49-2.53). Locus plots depicting the exact locations of the CNVs within these four regions are in Supplementary Figures S5–8. A cluster of four genes were found to locate at 19q13.32 including *OPA3, PPM1N, RTN2* and *VASP* intersecting with 168 long CNVs (Supplementary Figure S7). *PPM1N, RTN2* and *VASP* were also on the top signals and showed suggestive significance (Supplementary Tables S4-5). *CLCN6* at 1p36.22 and PSMB8 at 6p21.32 also showed suggestive significance for overall LC risk within the European ancestry population (Table 1). No duplications were found to be genome-wide significant in any scenario.

**Table 1.** Top significantly associated germline CNVs with lung cancer risk for gene-based association strategy (P-value < 5. 6 X 10^−6^). OR: odds ratio; CI: confidence interval; N of CNVs: mean number of CNVs in cases and controls; N of probes: mean number of probes covering these loci; All Lung: overall lung cancer risk in the whole population; EUR Lung: overall lung cancer risk in the European population. Demographic information including age, sex and smoking status, study site and four ancestry related PCs were adjusted in all regression models.

| Gene | Cytoband | Coordinates | N of CNVs | N of probes | Deletion |  | CNV<sup>a</sup> |  | Stratum |
| --- | --- | --- | --- | --- | --- | --- | --- | --- | --- |
|  |  |  |  |  | OR (95% CI) | P-value | OR (95% CI) | P-value |  |
| *CLCN6* | 1p36.22 | 11,866,248-11,903,187 | 265:133 | 24 | 1.82(1.45, 2.30) | $3.87 \times 10^{-7}$ | 1.79(1.42, 2.25) | $7.34 \times 10^{-7}$ | All Lung |
| *NFE2L2* | 2q31.2 | 178,095,031-178,129,859 | 160:73 | 16 | 2.01(1.48, 2.72) | $6.69 \times 10^{-6}$ | 2.03(1.50, 2.73) | $3.59 \times 10^{-6}$ | All Lung |
| *OPA3* | 19q13.32 | 46,030,685-46,088,060 | 105:36 | 22 | 2.63(1.76, 3.93) | $2.55 \times 10^{-6}$ | 2.63(1.76, 3.93) | $2.55 \times 10^{-6}$ | All Lung |
| *PSMB8*<sup>b</sup> | 6p21.32 | 32,808,494-32,814,277 | 210:97 | 50 | 1.94(1.49, 2.53) | $1.11 \times 10^{-6}$ | 1.89(1.45, 2.45) | $2.12 \times 10^{-6}$ | All Lung |
| *CLCN6* | 1p36.22 | 11,866,248-11,903,187 | 234:89 | 25 | 1.92(1.46, 2.52) | $2.62 \times 10^{-6}$ | 1.87(1.43, 2.45) | $4.95 \times 10^{-6}$ | EUR Lung |
| *PSMB8* | 6p21.32 | 32,808,494-32,814,277 | 196:70 | 50 | 2.07(1.53, 2.80) | $2.15 \times 10^{-6}$ | 2.03(1.51, 2.74) | $3.36 \times 10^{-6}$ | EUR Lung |

<sup>a</sup>The test was for the association between existence of CNVs (either duplications or deletions) and the risk of LC.
<sup>b</sup> CNVs from gene *PSMB* (Chr6:32,808,494-32,812,456) and *PSMB-AS1* (Chr6:32,811,863-32,814,277) were combined and labeled as *PSMB* here.

For the CNVR-based association, identified CNV calls were mapped to 8,892 CNV regions (CNVR), among which 314 were classified as common CNVR (frequency >1%), 6,905 and 1,673 were rare (frequency <1%) and singletons, respectively. Table 2 summarizes all significant findings which identified two genomic regions significantly associated with increased risk of LC, including 1p36.22 (Deletion OR=1.75, 95% CI=1.39-2.19; CNV OR=1.73, 95% CI=1.38-2.17), 2q31.2 (Deletion not reaching genome-wide significance level; CNV: OR=2.03, 95% CI=1.50-2.73). 22q12.3/22q13.1(Deletion OR=2.62, 95% CI=1.76-3.91, CNV OR=2.55, 95% CI=1.71-3.78) was also associated with increased susceptibility to lung adenocarcinoma (LUAD) in overall population.

**Table 2.** Top significantly associated germline CNV regions (CNVRs) with lung cancer risk (P-value < 5. 6 X 10^−6^). OR: odds ratio; 95% CI: 95% confidence interval; N of CNVs: mean number of CNVs in cases and controls; N of probes: mean number of probes covering these loci; All Lung: overall lung cancer risk in the whole population; All LUAD: adenocarcinoma risk in the whole population; EUR Lung: overall lung cancer risk in the European population. Demographic information including age, sex and smoking status, study site and four ancestry related PCs were adjusted in all regression models.

| CNVR |  |  |  |  | Deletion |  | CNV<sup>a</sup> |  |  |  |
| --- | --- | --- | --- | --- | --- | --- | --- | --- | --- | --- |
| ID | Cytoband | Coordinates | N of CNVs | N of probes | OR (95% CI) | P-value | OR (95% CI) | P-value | Gene list | Stratum<sup>b</sup> |
| CNVR_33 | 1p36.22 | 11,863,511-11,870,592 | 267:138 | 22 | 1.75(1.39, 2.19) | $1.68 \times 10^{-6}$ | 1.73(1.38, 2.17) | $2.10 \times 10^{-6}$ | *CLCN6, MTHFR* | All Lung |
| CNVR_1288 | 2q31.2 | 178,040,870-178,151,627 | 160:73 | 15 | 2.01(1.48, 2.72) | $6.69 \times 10^{-6}$ | 2.03(1.50, 2.73) | $3.59 \times 10^{-6}$ | *NFE2L2, HNRNPA3, LOC105373759, MIR4444-1, MIR4444-2, MIR3128, LOC100130691* | All Lung |
| CNVR_8852 | 22q12.3/22q13.1 | 37,578,488-37,686,988 | 64:51 | 17 | 2.62(1.76, 3.91) | $2.21 \times 10^{-6}$ | 2.55(1.71, 3.78) | $3.56 \times 10^{-6}$ | *C1QTNF6, CYTH4, SSTR3, RAC2* | All LUAD |

<sup>a</sup> The test was for the association between existence of duplications/deletions and the risk of lung cancer
<sup>b</sup> Results from subgroup analyses with sparse counts should be interpreted with caution and are considered exploratory.

Combining these results, we identified two highly confident CNV regions in 1p36.22 and 2q31.2 that were associated with LC susceptibility in overall population, among which the 1p36.22 region also showed suggestive significance for European population. The fact that these concordant results across both gene- and CNVR-based association analyses provides stronger evidence for their potential roles in LC. It is noted that the concordance evaluation of these results does not substitute for independent validation, but it may indicate that the signal is localized to the coding region rather than spanning broader regions. The number of probes covering these loci (Tables 1-2) and representative intensity plots (Supplementary Figure S3) provided initial evidence.

Stratified analyses by smoking status (e.g., ever vs. never smokers) for the top loci showed broadly consistent direction of effect estimates across strata, although statistical power was limited in never smokers due to small CNV counts (Supplementary Tables S6-7). This suggests that the observed associations were not solely driven by smoking exposure. Second, we examined whether CNV carrier status for the top loci was associated with smoking status (Supplementary Table S8). No associations were observed, indicating that the identified CNVs are not strongly correlated with smoking behavior.

### Sensitivity analyses showed preliminary findings of NFE2L2 in Asian population

To assess potential effect heterogeneity across genetic ancestries, the findings from our principal gene-based association analyses (Table 1) were further evaluated in the second-largest subpopulation in the TRICL consortium, the Asian population (n=2,826). Results of these sensitivity analyses are summarized in Supplementary Table S9. Notably, deletions or duplications in NFE2L2 was significantly associated with increased LC risk in the Asian population (Deletion OR=9.88, 95% CI=1.32-437.91, Fisher’s exact P-value=0.003; CNV OR=12.36, 95% CI=1.75-535.98, Fisher’s exact P-value=0.003). In parallel, these signals were not validated in the two independent validation cohorts (Supplementary Tables S10-11), which may be attributable to the limited sample sizes in both datasets and the small frequencies of germline CNVs in the general population. Also, the Affymetrix dataset used an Exome array with only about 100,000 genome-wide markers, so it can only detect larger CNVs. In particular, the CNVs of NFE2L2 at 2q31.2 showed a concordant direction of effect with a relatively large effect size and borderline statistical significance (OR = 3.59, 95% CI= [0.84, 15.29], p-value = 0.08) in the German OncoArray dataset, which is interpreted as suggestive evidence given the limited power (Supplementary Table S10).

### CNV-based polygenic risk score is associated with lung cancer risk

To further explore the cumulative contribution of these top variants, we conducted CNV-based PRS analyses using the significant signals identified from the TRICL discovery data (Table 1). The top four independent genes including *CLCN6, NFE2L2, OPA3* and *PSMB8* were used in the computation of PRS. All three PRS approaches (PRS weighted *PRS*_*β*_, unweighted *PRS*_*unw*_ and inverse variant weighted *PRS*_*IV*_) yielded significant associations with overall LC risk in the TRICL dataset (Table 3). The significance of the PRS was replicated in both validation cohorts: *PRS*_*β*_ showed significant association in the Germany OncoArray dataset (OR=1.19, P-value= 1.10 x 10^−3^) and the Affymetrix dataset (OR=1.06, P-value=0.03). Similarly, *PRS*_*IV*_ demonstrated significance in the Germany OncoArray dataset (OR=1.18, P-value=1.11 x 10^−3^). Among the three approaches, *PRS*_*β*_ consistently showed the strongest or equivalent associations (within *OR* ± 0.1) between the discovery and validation datasets.

**Table 3.** Association of polygenic risk score and lung cancer risk in the TRICL dataset for samples with European ancestry and two validation datasets. PRS: polygenic risk score; Validation set I: Germany OncoArray dataset; Validation set II: Affymetrix dataset; OR: odds ratio; CI: confidence interval. *PRS*_*β*_ : the standard PRS weights corresponding to the sum of the reported risk variant effect sizes (log (*β*)); *PRS*_*unw*_: unweighted sum of risk variants; *PRS*_*IV*_ : inverse variance weights that incorporate the standard error of the risk effect size. All these PRS have been standardized to have mean 0 and variance 1, so the ORs actually refer to increases of the PRS by one standard deviation.

|  |  | TRICL |  | Validation set I |  | Validation set II |  |
| --- | --- | --- | --- | --- | --- | --- | --- |
|  |  | OR (95% CI) | P-value | OR (95% CI) | P-value | OR (95% CI) | P-value |
| **Top genes<sup>a</sup>** | $PRS_{\beta}$ | 1.08(1.05, 1.11) | $9.32 \times 10^{-9}$ | 1.19(1.07, 1.32) | $1.10 \times 10^{-3}$ | 1.06(1.00, 1.12) | **0.03** |
| | $PRS_{unw}$ | 1.08(1.05, 1.11) | $1.05 \times 10^{-8}$ | 0.93(0.85, 1.02) | 0.14 | 1.05(0.99, 1.11) | 0.08 |
| | $PRS_{IV}$ | 1.08(1.05, 1.11) | $1.04 \times 10^{-8}$ | 1.18(1.07, 1.30) | $1.11 \times 10^{-3}$ | 1.05(1.00, 1.11) | 0.07 |
| **Top 20 genes<sup>b</sup>** | $PRS_{\beta}$ | 1.09(1.06, 1.12) | $1.29 \times 10^{-9}$ | 1.22(1.10, 1.36) | $1.94 \times 10^{-4}$ | 1.06(1.00, 1.12) | **0.03** |
| | $PRS_{unw}$ | 1.09(1.06, 1.12) | $2.27 \times 10^{-9}$ | 0.96(0.87, 1.06) | 0.41 | 1.05(1.00, 1.11) | 0.06 |
| | $PRS_{IV}$ | 1.09(1.06, 1.12) | $2.02 \times 10^{-9}$ | 1.20(1.09, 1.32) | $3.47 \times 10^{-4}$ | 1.05(1.00, 1.11) | 0.06 |

<sup>a</sup>Top significant genes from the TRICL gene-based association analysis
<sup>b</sup>Top 20 significant genes from the TRICL gene-based association analysis

To explore the contribution of additional loci, we further constructed PRS models using an expanded set of the top twenty signals ranked by association strength (Supplementary Table S12). The comparable performance between the original PRS model (i.e., only four genes) and expanded models (i.e., top 20 genes) suggests that the strongest signals capture the majority of the CNV-related genetic risk for lung cancer (Table 3). Our PRS performance may appear over-optimistic as we used the estimated effect sizes from each dataset to construct PRS, which may introduce overfitting and inflate association estimates. Therefore, these results should be interpreted as internal validation instead of external replication.

### Pathway Enrichment Analyses

We further explored the biological pathways associated with the candidate genes to gain insights into their potential functional impact. Based on the top 200 significant genes, gene set enrichment analysis identified six significant KEGG pathways (i.e., FDR<0.05, Supplementary Table S13), including three cancer-related pathways: acute myeloid leukemia, chronic myeloid leukemia, and pathways in cancer; along with one apoptosis-related pathway (apoptosis) and one immune-related pathway, Fc gamma receptor (FcγR)-mediated phagocytosis (Supplementary Figure S9, Supplementary Table S13). Consistently, 11 LC-associated pathways were also identified through hallmark pathways, which highlighted oncogenic signaling (TNF⍰signaling via NF-*k*B and MYC targets), apoptosis-related processes (apoptosis, ultraviolet response, oxidative phosphorylation, G_2_M checkpoint), and immune response pathways (allograft rejection, interferon gamma response, inflammatory response, and interleukin-2/STAT5 signaling) (Supplementary Figure S9). Notably, two candidate genes identified in our CNV analysis, *NFE2L2* and *PSMB8*, were directly involved in key immune and inflammatory signal pathways, including TNF⍰ signaling via NF-*k*B and the interferon gamma response, respectively. These findings indicate that these genetic alterations may contribute to cancer risk through multiple mechanisms, including effects on inflammation, cell survival and proliferation, and apoptosis (43,44). Importantly, these pathways are closely linked to deficiencies in DNA damage and repair (45,46), which represent critical precursors to genome instability—a defining hallmark of cancer (25,47).

### Functional findings are consistent with a role for NFE2L2 and OPA3 in genomic stability

To assess the functional relevance of the identified LC susceptibility CNVs, we performed gene knockdown experiments targeting the identified four regions in a lung fibroblast cell line (Table 1). *PPM1N, RTN2* and *VASP* largely intersecting with *OPA3* at 19q13.32 were also included in the evaluation. Among the tested genes, silencing *NFE2L2* or *OPA3* was associated with increased DNA damage, as indicated by elevated γH2AX levels (unpaired two-tailed t-test *P*-values: 0.0255 for *NFE2L2* and 0.0235 for *OPA3*) (Figure 3A, left panel).

**Fig. 3.**
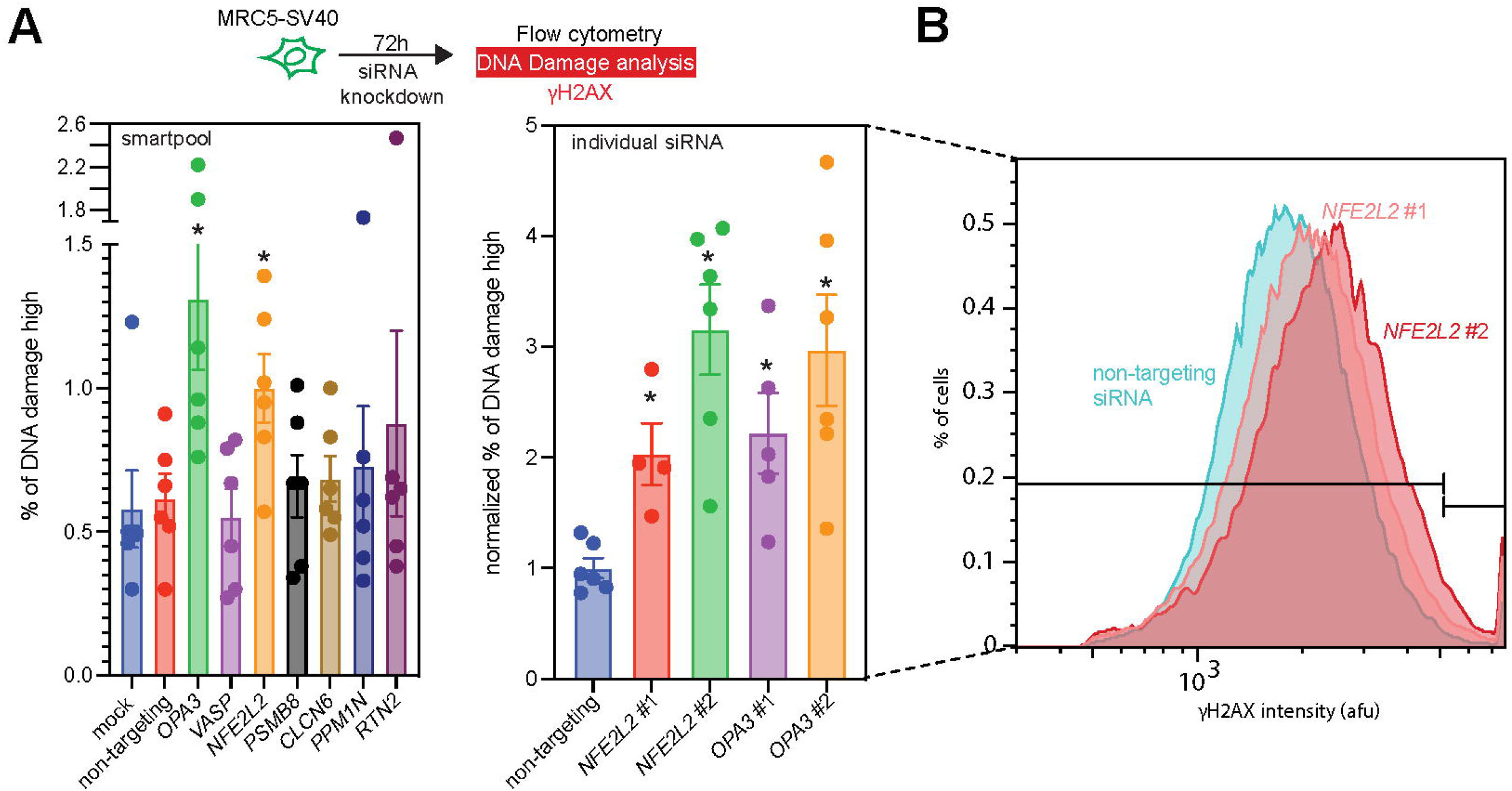
Analysis of DNA Damage Induced by siRNA Knockdown. (A) left: Bar graph illustrating the percentage of high DNA damage (γH2AX) signal, a marker for DNA double-strand breaks, at 72 hours post-transfection with either SMARTpool or individual siRNAs fold increase in γH2AX signal compared to non-targeting control siRNA. Each bar corresponds to a specific gene, with colors representing different siRNA treatments. Error bars indicate standard error. Data points marked with an asterisk (*) denote statistical significance as determined by a two-tailed unpaired t-test. (B) Representative flow cytometry histogram of γH2AX intensity in cells transfected with two different siRNAs targeting *NFE2L2* (NFE2L2 #1 and NFE2L2 #2) compared to a non-targeting siRNA control. The x-axis displays γH2AX intensity (arbitrary units), and the y-axis shows the frequency of cells. The shift in the histogram to the right for *NFE2L2* siRNAs indicates an increase in γH2AX signal, thus higher DNA damage, compared to the control.

To mitigate potential off-target effects, we performed follow-up experiments using individual siRNAs targeting each gene. These experiments yielded consistent results, with increased γH2AX levels observed following knockdown of *NFE2L2* (P = 0.003 and 0.0004 for siRNAs #1 and #2) and *OPA3* (P = 0.006 and 0.003 for siRNAs #1 and #2) (Figure 3A, right panel). Representative flow cytometry histograms (Figure 3B) further supported these findings, showing increased γH2AX signal across independent siRNAs.

Taken together, these results are consistent with a role for NFE2L2 and OPA3 in maintaining genomic integrity in human lung fibroblasts. However, these experiments were conducted in fibroblasts rather than epithelial cells, which are more directly implicated in lung carcinogenesis, and the reported P-values are nominal without formal multiple comparison adjustment across genes and siRNAs. Accordingly, these findings should be considered exploratory and supportive. Within this context, our results suggest that CNV-driven disruption of NFE2L2 and OPA3 may contribute to genomic instability, a hallmark of cancer, providing a basis for further investigation in more disease-relevant models.

## Discussion

In this study, we conducted one of the largest genome-wide investigations of germline CNVs in relation to LC, leveraging data from TRICL and two independent validation cohorts. We identified four genomic regions, 1p36.22, 2q31.2, 6p21.32, and 19q13.32, associated with increased LC risk, with two loci (1p36.22 and 2q31.2) supported by both gene-based and CNV region–based analyses. These findings extend prior SNP-based genome-wide association studies by implicating structural variation as an additional contributor to inherited susceptibility. We did not observe substantial heterogeneity across histological subtypes, although this may reflect limited statistical power for subtype-specific analyses. Functional evaluation further suggests that disruption of *NFE2L2* (2q31.2) and *OPA3* (19q13.32) may promote genomic instability, supporting their potential relevance in LC carcinogenesis.

Among the two high-confidence regions, 1p36.22 has been previously implicated in other carcinoma (48) and the broader 1p36 genomic region is known to harbor tumor suppressor genes (49). Similarly, 2q31.2 has been reported as a susceptibility locus in other cancers (50). Notably, this region includes NFE2L2, a key regulator of oxidative stress response, for which copy number gains and activating mutations have been observed in lung tumors (28-30). Our findings suggest that, beyond somatic alterations, inherited CNVs affecting these regions may predispose individuals to LC. Disruption of NFE2L2 may influence carcinogenesis and downstream DNA damage responses. In addition, our analysis also highlighted 6p21.32 which are consistent with prior reports identifying 6p21.32 in both European and Asian populations (26,27). Interestingly, locus plots of the identified CNVs on 6p21.32 from our study are located in different regions compared with those from previous reports (26,27), suggesting a potential previously unrecognized mechanism of disease susceptibility of LC.

We also identified a cluster of genes at 19q13.32 including OPA3, PPM1N, RTN2, and VASP. This region has been implicated in prior LC genetic studies (51,52), suggesting potential overlap with known susceptibility loci. Among these genes, VASP has been shown to be upregulated in lung adenocarcinoma and to promote tumor cell migration and invasion (53-55), supporting its potential role in tumor progression. The co-localization of multiple genes within the same CNV region raises the possibility that coordinated dosage alterations may collectively influence cancer-related pathways. However, the individual and combined functional roles of these genes require further investigation.

Despite these strengths, several limitations should be noted. First, the principal CNV and LC associations from the TRICL discovery dataset did not replicate convincingly in the validation cohorts. Although harmonization of the CNV definition was achieved, differences in platform probe density and CNV detectability likely contributed to reduced comparability across datasets. In addition, the low frequency of germline CNVs observed in our study substantially limits statistical power in the validation cohorts and sensitivity analyses. As such, variability in effect estimates including occasional directional inconsistency is expected and should therefore be interpreted as exploratory. We further emphasize that concordance between gene-based and CNVR-based analyses within the discovery dataset provides internal consistency and supports the robustness of signal localization.

Another consideration is the pooling strategy across ancestries in the TRICL discovery data overall analyses, which is supported by recent literature (56). While cross-ancestry pooling increases power, particularly for rare CNV detection, it may introduce residual population stratification despite adjustment using principal components. Nevertheless, the concordance of results in ancestry-stratified analyses supports the robustness of our findings. A third limitation is the use of a lung fibroblast model rather than epithelial cells for functional evaluation, given that epithelial cells are central to lung adenocarcinoma pathogenesis. While this system provides a robust platform to study DNA damage and early genomic instability, future studies in epithelial and other lung cell types will be important to confirm and extend these findings.

From a translational and population health perspective, our findings suggest that germline CNVs may contribute to risk stratification in cancer susceptibility. A subset of the identified CNVs could potentially be incorporated into targeted screening panels for large-scale population-based studies. In addition, the developed CNV-based polygenic risk score (CNV-PRS) may complement existing risk prediction and enable more refined stratification in future screening and prevention strategies. However, further validation in independent and ethnically diverse cohorts will be necessary before these findings can be translated into routine practice. Future studies with larger sample sizes and harmonized CNV detection approaches will be essential to enable robust evaluation of subgroup-specific effects (e.g., by sex and age) and to more rigorously assess gene–environment (e.g., genetic and smoking) interactions. In addition, the integration with transcriptomic and network-based analyses will be important to evaluate potential dose–response relationships, uncover novel biological pathways, and further provide mechanistic insights into CNV-associated LC risk.

## Acknowledgements

Feifei Xiao was supported by NIH/NHGRI R21HG010925. Jun Xia was supported by Texas A&M startup fund and NIH/NIEHS R00ES033259 awards. This project was also supported by the Cytometry and Cell Sorting Core at Baylor College of Medicine with funding from the CPRIT Core Facility Support Award (CPRIT-RP180672), the NIH (P30 CA125123 and S10 RR024574) and the expert assistance of Joel M. Sederstrom.

## Data Availability

The Lung OncoArray Project is available in the database for Phenotypes and Genotypes (dbGAP) as phs001273.v4.p2. The Affymetrix Axiom Array from the Transdisciplinary Research into Cancer of the Lung project is available in dbGAP as phs001681.v1.p1. Data from the validation GWAS using the OncoArray has been uploaded as a part of the Transdisciplinary Research into Cancer of the Lung (TRICL): CIDR Lung Cancer Whole Exome Sequencing Project, phs000878.v2.p1.

## Author Contributions

Drafting of the manuscript: Xiao F, Fei Qin, Amos CI. Project Coordination: Xiao F, Amos CI. CNV calling and association analysis: Qin F, Luo X, Xiao F, Amos CI. Polygenetic risk score analysis: Luo X, Qin F, Xiao F. Pathway analysis: Qin F, Cai G. Functional experiment to evaluate the impact of seven top significant genes in lung cell line: Xia J, Slewitzke SE, Fernandes GF.

