## Supplementary Tables and Figures for "Large-scale association study identifies lung cancer susceptibility copy number variants and their potential functional role in genetic instability"

**Supplementary Table S1. Demographic characteristics of the TRICL samples after genotyping quality control filters.**

|  | **Lung cancer cases**  **(n=18,455)** | | **Controls**  **(n=15,065)** | |
| --- | --- | --- | --- | --- |
|  | **Number** | **%** | **Number** | **%** |
| **Age** |  |  |  |  |
| <=50 years | 1,419 | 8 | 1,696 | 11 |
| >50 years | 17,036 | 92 | 13,369 | 89 |
| **Sex** |  |  |  |  |
| Male | 11,558 | 62 | 9,312 | 61 |
| Female | 6,872 | 37 | 5,738 | 39 |
| **Smoking status** |  |  |  |  |
| Never | 2,118 | 11 | 4,868 | 32 |
| Ever | 682 | 4 | 721 | 5 |
| Former | 6,346 | 34 | 4,412 | 29 |
| Current | 9,032 | 49 | 4,767 | 32 |
| **Histology**^a^ |  |  |  |  |
| Adenocarcinoma | 7,544 | 41 | - |  |
| Squamous cell carcinoma | 4,468 | 24 | - |  |
| Small cell carcinoma | 1,719 | 9 | - |  |
| **Race** |  |  |  |  |
| European | 16,556 | 90 | 12,768 | 85 |
| Asians | 1,268 | 7 | 1,558 | 10 |
| African Americans | 427 | 2 | 545 | 4 |
| Others | 204 | 1 | 194 | 1 |

^a^The remaining 25.7% include other histological subsets, such as large cell carcinoma, non-small cell lung cancer, mixed histology and unknown.

**Supplementary Table S2. Summary of the CNV quality visual inspection for modSaRa2 and PennCNV.** CNV calls from PennCNV and modSaRa2 were examined by visualization of the signal intensities. 400~800 calls were random selected for each scenario in the visualization. The power of these two methods was measured in the overlapping and non-overlapping CNVs separately. For the overlapping ones, three thresholds were used in proportion of overlapping CNVs by length, including >0%, >20% and >50%, respectively. The true positive rates (TPRs) were evaluated under scenarios of different copy number states (i.e., duplications and deletions). For non-overlapping ones, the TPR was calculated for different CNV lengths (< 50, 50-200 and > 200kb). The results suggested merging the CNV sets from two callers. We substituted the CNV calls > 200kb detected by PennCNV with those from modSaRa2.

| **TPR** | **overlapping** | | | | | | **non-overlapping** | | | | | |
| --- | --- | --- | --- | --- | --- | --- | --- | --- | --- | --- | --- | --- |
|  | **Deletion** | | | **Duplication** | | | **Deletion** | | | **Duplication** | | |
|  | **0%** | **20%** | **50%** | **0%** | **20%** | **50%** | **<50** | **50~200** | **>200** | **<50** | **50~200** | **>200** |
| **modSaRa2** | 0.86 | 0.91 | 0.95 | 0.89 | 0.91 | 0.92 | 0.96 | 0.96 | **0.95** | 0.63 | 0.66 | ~~--~~ |
| **PennCNV** | 0.90 | 0.94 | 0.96 | 0.88 | 0.95 | 0.96 | 0.99 | 0.95 | **0.87** | 0.97 | 1.00 | 0.95 |

**Supplementary Table S3. Summary of the merged CNV calling results from PennCNV and modSaRa2.** The number of deletions and duplications in cases and controls for all samples (including all races). The total number of CNVs in each sample and the size (kb) distribution of CNVs are listed in mean and median.

| **Status** | **N** | **N del** | **N dup** | **Mean N of CNVs** | **Median N of CNVs** | **Mean size (kb)** | **Median size (kb)** |
| --- | --- | --- | --- | --- | --- | --- | --- |
| **Cases** | 18,455 | 542,043 | 133,949 | 36.63 | 23 | 37.30 | 11.49 |
| **Control** | 15,065 | 458,813 | 108,530 | 37.66 | 23 | 37.07 | 11.89 |
| **Total** | 33,520 | 1,000,856 | 242,479 | 37.09 | 23 | 37.20 | 11.70 |

N: number of samples; N del: number of deletions; N dup: number of duplications; mean N of CNV: mean number of CNVs per sample; median N of CNVs: median number of CNVs.

**Supplementary Table S4. Full set of association results from the gene-based association approach.** The results for overall population, by histological subtypes, ancestry subsets, by deletion or duplication or any CNV are provided. For simplicity, we only reported the genes with OR>1 in the main study report to show the significant risk CNVs instead of protective ones.

*Tables are in separate excel worksheet.*

**Supplementary Table S5. Full set of association results from the CNVR association approach.** The results for overall population, by histological subtypes, ancestry subsets, by deletion or duplication or any CNV are provided. For simplicity, we only reported the genes with OR>1 in the main study report to show the significant risk CNVs instead of protective ones.

*Tables are in separate excel worksheet.*

**Supplementary Table S6: Stratified association analyses by smoking status (e.g., ever vs. never smokers) for the top loci identified in the primary analyses.** The test was for the association between existence of CNVs (either duplications or deletions) and the risk of lung cancer. OR: odds ratio; 95% CI: 95% confidence interval; N of CNVs: mean number of CNVs in cases and controls; All Lung: overall lung cancer risk in the whole population; Demographic information including age, sex, study site and four ancestry related PCs were adjusted in all regression models. Note that these p-values are nominal, given the global p-value threshold (5.60 $\times{10}^{-6}$), only *CLCN6* showed genome wide significance in smokers.

|  |  | **Smokers (N=25,960)** | | | **Non-smokers (N=6,986)** | | |  |
| --- | --- | --- | --- | --- | --- | --- | --- | --- |
| **Gene** | **Cytoband** | **N of CNVs** | **OR (95% CI)** | **P-value** | **N of CNVs** | **OR (95% CI)** | **P-value** | **Stratum**^a^ |
| ***CLCN6*** | 1p36.22 | 236:105 | 1.82(1.42, 2.34) | **1.87** $\boldsymbol{\times}\boldsymbol{10}^{\boldsymbol{-6}}$ | 27:27 | 1.26(0.70, 2.29) | 0.44 | All Lung |
| ***NFE2L2*** | 2q31.2 | 137:61 | 1.82(1.32, 2.50) | 2.28$\times{10}^{-4}$ | 23:12 | 2.70(1.22, 5.93) | 0.01 | All Lung |
| ***OPA3*** | 19q13.32 | 90:28 | 2.42(1.56, 3.75) | 7.54 $\times{10}^{-5}$ | 15:8 | 1.95(0.76, 4.99) | 0.17 | All Lung |
| ***PSMB8*** | 6p21.32 | 186:82 | 1.79(1.36, 2.37) | 3.63 $\times{10}^{-5}$ | 24:15 | 1.84(0.89, 3.82) | 0.10 | All Lung |

^a^ Results from subgroup analyses with sparse counts should be interpreted with caution and are considered exploratory.

**Supplementary Table S7: Stratified association analyses by smoking** **status (e.g., ever vs. never smokers) for the top loci identified in the primary analyses.** The test was for the association between deletions and the risk of lung cancer. OR: odds ratio; 95% CI: 95% confidence interval; N of CNVs: mean number of CNVs in cases and controls; All Lung: overall lung cancer risk in the whole population. Demographic information including age, sex, study site and four ancestry related PCs were adjusted in all regression models. Note that these p-values are nominal, given the global p-value threshold (5.60 $\times{10}^{-6}$), only *CLCN6* showed genome wide significance in smokers.

|  |  | **Smokers (N=25,960)** | | | **Non-smokers (N=6,986)** | | |  |
| --- | --- | --- | --- | --- | --- | --- | --- | --- |
| **Gene** | **Cytoband** | **N of CNVs** | **OR (95% CI)** | **P-value** | **N of CNVs** | **OR (95% CI)** | **P-value** | **Stratum**^a^ |
| ***CLCN6*** | 1p36.22 | 236:103 | 1.87(1.46, 2.39) | **8.78** $\boldsymbol{\times}\boldsymbol{10}^{\boldsymbol{-7}}$ | 26:26 | 1.25(0.68, 2.29) | 0.48 | All Lung |
| ***NFE2L2*** | 2q31.2 | 135:61 | 1.76(1.28, 2.43) | 4.70$\times{10}^{-4}$ | 22:9 | 3.27(1.37, 7.81) | 7.45 $\times{10}^{-3}$ | All Lung |
| ***OPA3*** | 19q13.32 | 90:28 | 2.42(1.56, 3.75) | 7.54 $\times{10}^{-5}$ | 15:8 | 1.95(0.76, 4.99) | 0.17 | All Lung |
| ***PSMB8*** | 6p21.32 | 185:80 | 1.80(1.36, 2.38) | 3.53 $\times{10}^{-5}$ | 24:13 | 2.04(0.96, 4.34) | 0.06 | All Lung |

^a^ Results from subgroup analyses with sparse counts should be interpreted with caution and are considered exploratory.

**Supplementary Table S8: Association between smoking status (e.g., ever vs. never smokers) and the top loci identified in the primary analyses.** The test was for the association between deletions and the risk of lung cancer. OR: odds ratio; 95% CI: 95% confidence interval; N of CNVs: mean number of CNVs in smokers and non-smokers; All Lung: overall lung cancer risk in the whole population. Adjusted covariates including age, sex, study site, case-control status and four ancestry related PCs were adjusted in regression models. Note that these p-values are nominal, given the global p-value threshold (5.60 $\times{10}^{-6}$), none showed genome wide significance.

|  |  | **CNV** | | | **Deletion** | | |  |
| --- | --- | --- | --- | --- | --- | --- | --- | --- |
| **Gene** | **Cytoband** | **N of CNVs** | **OR (95% CI)** | **P-value** | **N of CNVs** | **OR (95% CI)** | **P-value** | **Stratum** |
| ***CLCN6*** | 1p36.22 | 341:57 | 0.72(0.52, 0.99) | 0.04 | 339:55 | 0.73(0.52, 1.01) | 0.06 | All Lung |
| ***NFE2L2*** | 2q31.2 | 198:35 | 0.56(0.37, 0.84) | 5.02 $\times{10}^{-3}$ | 196:31 | 0.60(0.39, 0.93) | 0.02 | All Lung |
| ***OPA3*** | 19q13.32 | 118:23 | 0.44(0.26, 0.73) | 1.48 $\times{10}^{-3}$ | 118:23 | 0.44(0.26, 0.73) | 1.48 $\times{10}^{-3}$ | All Lung |
| ***PSMB8*** | 6p21.32 | 268:39 | 0.65(0.44, 0.95) | 0.03 | 265:37 | 0.65(0.44, 0.96) | 0.03 | All Lung |

**Supplementary Table S9. Sensitivity analysis of top significant genes in the Chinese population.** OR: odds ratio computed from the Wald test; CI: confidence interval; P-value: P-values from Wald test; Exact P-value: P-value from Fisher’s exact test. Results from these analyses with sparse counts should be interpreted with caution and are considered exploratory.

|  |  |  | | **Deletion** | | | | **CNV^a^** | | |
| --- | --- | --- | --- | --- | --- | --- | --- | --- | --- | --- |
| **Gene** | **Cytoband** | **N of CNVs**  **in Cases: Controls** | **OR (95% CI)** | | **P-value** | **Exact P-value** | **OR (95% CI)** | | **P-value** | **Exact P-value** |
| ***CLCN6*** | 1p36.22 | 19:25 | 1.20(0.63, 2.29) | | 0.57 | 0.88 | 1.20(0.63, 2.29) | | 0.57 | 0.88 |
| ***NFE2L2*** | 2q31.2 | **9:0** | **9.88(1.32, 437.91)^b^** | | **-** | **0.01** | **12.36(1.75, 535.98)^b^** | | **-** | **0.003** |
| ***OPA3*** | 19q13.32 | 1:3 | 0.26(0.02, 2.89) | | 0.27 | 0.63 | 0.26(0.02, 2.89) | | 0.27 | 0.63 |
| ***PPM1N*** | 19q13.32 | 2:3 | 0.55(0.08, 3.86) | | 0.55 | 1.00 | 0.55(0.08, 3.86) | | 0.55 | 1.00 |
| ***PSMB8*** | 6p21.32 | 6:5 | 1.72(0.37, 8.07) | | 0.49 | 0.48 | 1.40(0.39, 4.95) | | 0.61 | 0.56 |
| ***RTN2*** | 19q13.32 | 2:3 | 0.55(0.08, 3.86) | | 0.55 | 1.00 | 0.55(0.08, 3.86) | | 0.55 | 1.00 |
| ***VASP*** | 19q13.32 | 2:3 | 0.55(0.08, 3.86) | | 0.55 | 1.00 | 0.55(0.08, 3.86) | | 0.55 | 1.00 |

^a^ The test was for the association between existence of CNVs (either duplications or deletions) and the risk of LC

^b^ The odds ratios were computed from the Fisher’s exact test

**Supplementary Table S10. Validation analysis of top significant genes in German Oncoarray dataset (n=1,851).** Gene-based strategy was used in this validation analysis. N of CNVs: mean number of CNVs in cases and controls; OR: odds ratio; CI: 9confidence interval; P-value: P-values from Wald test; Exact P-value: P-value from Fisher’s exact test.

|  |  |  | **Deletion** | | | **CNV^a^** | | |
| --- | --- | --- | --- | --- | --- | --- | --- | --- |
| **Gene** | **Cytoband** | **N of CNVs** | **OR (95% CI)** | **P-value** | **Exact**  **P-value** | **OR (95% CI)** | **P-value** | **Exact**  **P-value** |
| ***CLCN6*** | 1p36.22 | 3:7 | 0.20(0.04, 0.94) | 0.04 | 0.07 | 0.16(0.04, 0.71) | 0.02 | 0.04 |
| ***NFE2L2*** | 2q31.2 | 9:3 | 3.59(0.84, 15.29) | 0.08 | 0.56 | 3.59(0.84, 15.29) | 0.08 | 0.56 |
| ***OPA3*** | 19q13.32 | 2:3 | 0.43(0.07, 2.73) | 0.37 | 0.35 | 0.43(0.07, 2.73) | 0.37 | 0.35 |
| ***PPM1N*** | 19q13.32 | 4:5 | 0.37(0.09, 1.46) | 0.16 | 0.29 | 0.37(0.09, 1.46) | 0.16 | 0.29 |
| ***PSMB8*** | 6p21.32 | 6:4 | 0.58(0.27, 1.23) | 0.15 | 0.11 | 0.58(0.27, 1.23) | 0.15 | 0.11 |
| ***RTN2*** | 19q13.32 | 4:5 | 0.37(0.09, 1.46) | 0.16 | 0.29 | 0.37(0.09, 1.46) | 0.16 | 0.29 |
| ***VASP*** | 19q13.32 | 4:5 | 0.37(0.09, 1.46) | 0.16 | 0.29 | 0.37(0.09, 1.46) | 0.16 | 0.29 |

^a^ The test was for the association between existence of CNVs (either duplications or deletions) and the risk of LC

**Supplementary Table S11. Validation analysis of top significant genes in the Affymetrix dataset (n=6,035).** Gene-based strategy was used in this validation analysis. N of CNVs: mean number of CNVs in cases and controls; OR: odds ratio; CI: confidence interval; P-value: P-values from Wald test; Exact P-value: P-value from Fisher’s exact test.

|  |  |  | **Deletion** | | | **CNV**^a^ | | |
| --- | --- | --- | --- | --- | --- | --- | --- | --- |
| **Gene** | **Cytoband** | **N of CNVs** | **OR (95% CI)** | **P-value** | **Exact P-value** | **OR (95% CI)** | **P-value** | **Exact**  **P-value** |
| ***CLCN6*** | 1p36.22 | 241:357 | 1.13(0.94, 1.35) | 0.20 | - | 1.13(0.94, 1.35) | 0.19 | - |
| ***NFE2L2*** | 2q31.2 | 2:1 | 4.15(0.37, 46.01) | 0.25 | 0.57 | 4.15(0.37, 46.01) | 0.25 | 0.57 |
| ***PSMB8*** | 6p21.32 | 31:40 | 1.48(0.91, 2.42) | 0.12 | - | 1.48(0.91, 2.42) | 0.12 | - |

^a^ The test was for the association between existence of CNVs (either duplications or deletions) and the risk of LC

**Supplementary Table S12. Top significant genes used for calculation of polygenic risk score and its association with lung cancer risk in the TRICL dataset.** OR: odds ratio; CI: confidence interval; SIG: significant genes from TRICL gene-based association analysis (P-value< $5.6\times{10}^{-6}$); Top 20: top 20 significant genes from TRICL gene-based association analysis. For genes on the same genomic band, the one with smallest P-value were remained in calculating polygenic risk score.

| **Id** | **Genes** | **Coordinate** | **OR (95% CI)** | **P-value** | **SIG** | **Top 20** |
| --- | --- | --- | --- | --- | --- | --- |
| 1 | CLCN6 | 1p36.22 | 1.79(1.42, 2.25) | 7.34 $\times{10}^{-7}$ | 🗸 | 🗸 |
| 2 | PSMB8-AS1^a^ | 6p21.32 | 1.89(1.45, 2.45) | 2.12 $\times{10}^{-6}$ | 🗸 | 🗸 |
| 3 | OPA3 | 19q13.32 | 2.63(1.76, 3.93) | 2.55 $\times{10}^{-6}$ | 🗸 | 🗸 |
| 4 | NFE2L2 | 2q31.2 | 2.03(1.50, 2.73) | 3.59 $\times{10}^{-6}$ | 🗸 | 🗸 |
| 5 | PPM1N | 19q13.32 | 2.25(1.57, 3.22) | 9.29 $\times{10}^{-6}$ | - | - |
| 6 | RTN2 | 19q13.32 | 2.25(1.57, 3.22) | 9.29 $\times{10}^{-6}$ | - | - |
| 7 | VASP | 19q13.32 | 2.26(1.58, 3.24) | 9.45 $\times{10}^{-6}$ | - | - |
| 8 | PSMB8 | 6p21.32 | 1.82(1.39, 2.36) | 9.52 $\times{10}^{-6}$ | - | - |
| 9 | C16orf72 | 16p13.2 | 2.46(1.64, 3.70) | 1.47 $\times{10}^{-5}$ |  | 🗸 |
| 10 | TREM1 | 6p21.1 | 2.79(1.72, 4.51) | 2.93 $\times{10}^{-5}$ |  | 🗸 |
| 11 | FKBP8 | 19p13.11 | 3.30(1.88, 5.81) | 3.48 $\times{10}^{-5}$ |  | 🗸 |
| 12 | PRKCH | 14q23.1 | 2.07(1.46, 2.95) | 5.06 $\times{10}^{-5}$ |  | 🗸 |
| 13 | IL1RN | 2q14.1 | 2.34(1.55, 3.52) | 5.06 $\times{10}^{-5}$ |  | 🗸 |
| 14 | CYP27A1 | 2q35 | 3.53(1.91, 6.53) | 5.56 $\times{10}^{-5}$ |  | 🗸 |
| 15 | SPEN | 1p36.21-p36.13 | 3.63(1.92, 6.88) | 7.61 $\times{10}^{-6}$ |  | 🗸 |
| 16 | LST1 | 6p21.33 | 2.07(1.44, 2.98) | 8.51 $\times{10}^{-6}$ |  | 🗸 |
| 17 | MTHFS | 15q25.1 | 3.59(1.90, 6.80) | 8.54 $\times{10}^{-6}$ |  | 🗸 |
| 18 | SUCNR1 | 3q25.1 | 4.00(2.00, 7.99) | 8.66 $\times{10}^{-5}$ |  | 🗸 |
| 19 | NEDD9 | 6p24.2 | 2.26(1.50, 3.40) | 8.67 $\times{10}^{-5}$ |  | 🗸 |
| 20 | FLJ22447 | 14q23.1-q23.2 | 2.24(1.49, 3.34) | 9.05 $\times{10}^{-5}$ |  | 🗸 |

^a^ PSMB8-AS1 and PSMB8 are the same gene region. PSMB-AS1 is the long non-coding RNA transcribed in the antisense orientation relative to PSMB8. We merged these two genes in all downstream analyses.

**Supplementary Table S13. Summary of significant pathways and involved gene lists.** Gene set enrichment analysis (GSEA) was performed on the top 200 CNV-associated genes in the gene-based and CNVR-based association analyses. Enrichment was tested against KEGG and MsigDB hallmark pathways. P-values and gene list for each pathway from the Hallmark and KEGG analyses are summarized.

*Tables are in separate excel worksheet*

**Supplementary Figure S1. Overview of PennCNV and modSaRa2 CNV calling and quality control.** The figure outlines the study design of CNV calling including a brief description of quality control steps. Summary of key results includes the sample sizes and number of CNVs at various stages of analysis. IBD: identity by descent; LRR_SD: standard deviation of log R ratio on chromosome 1.

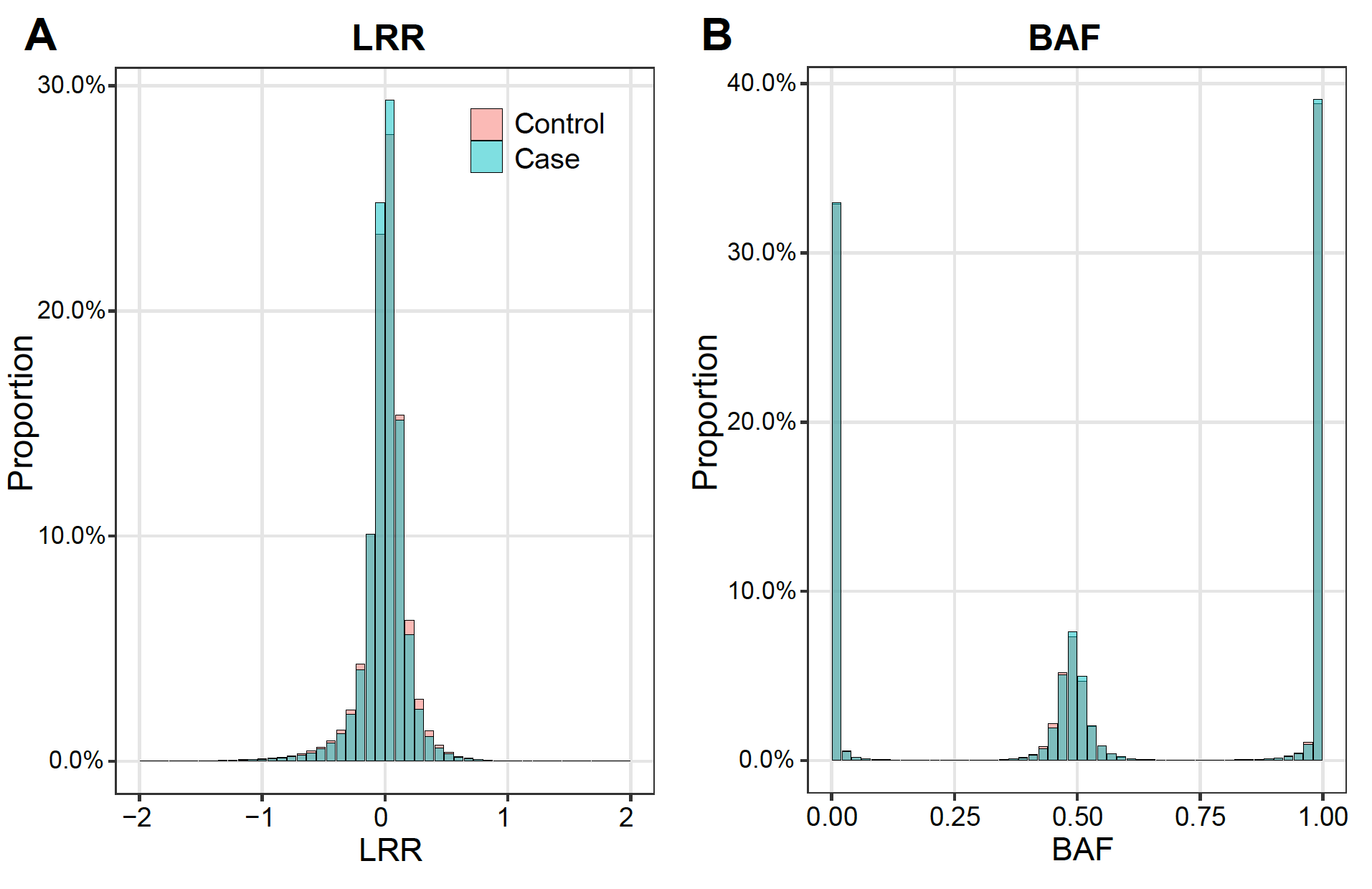

**Supplementary Figure S2. Distribution of Log R Ratio and B allele frequency in TRICL samples.** The distributions of LRR and BAF for chromosomes 1 and 2 across 500 representative samples (289 cases and 211 controls). Overall, the LRR and BAF distributions are highly similar between cases and controls. LRR: Log R Ratio; BAF: B allele frequency.

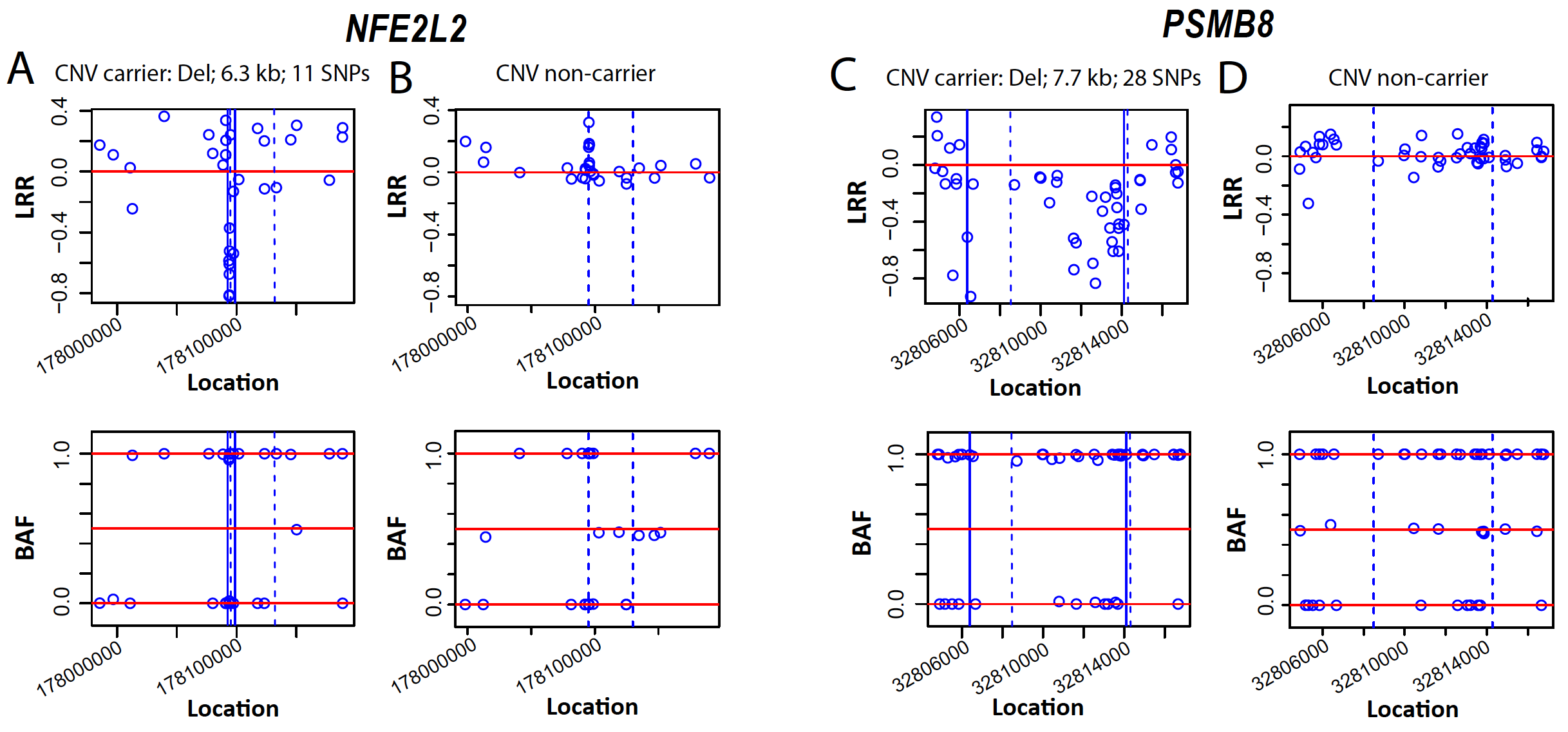

**Supplementary Figure S3**: **LRR and BAF profiles for two representative genes in CNV carriers and non-carriers.** The top panels show LRR values, and the bottom panels show BAF values. Panels (A) and (B) correspond to CNV carrier and non-carrier samples for *NFE2L2*, respectively, while panels (C) and (D) correspond to CNV carrier and non-carrier samples for *PSMB8*. Dashed blue vertical lines indicate the genomic locations of the genes, and solid blue vertical lines mark the boundaries of the CNVs. For CNV carriers, the CNV type, width, and number of probes within the CNV are displayed at the top of each plot. LRR: Log R Ratio; BAF: B allele frequency.

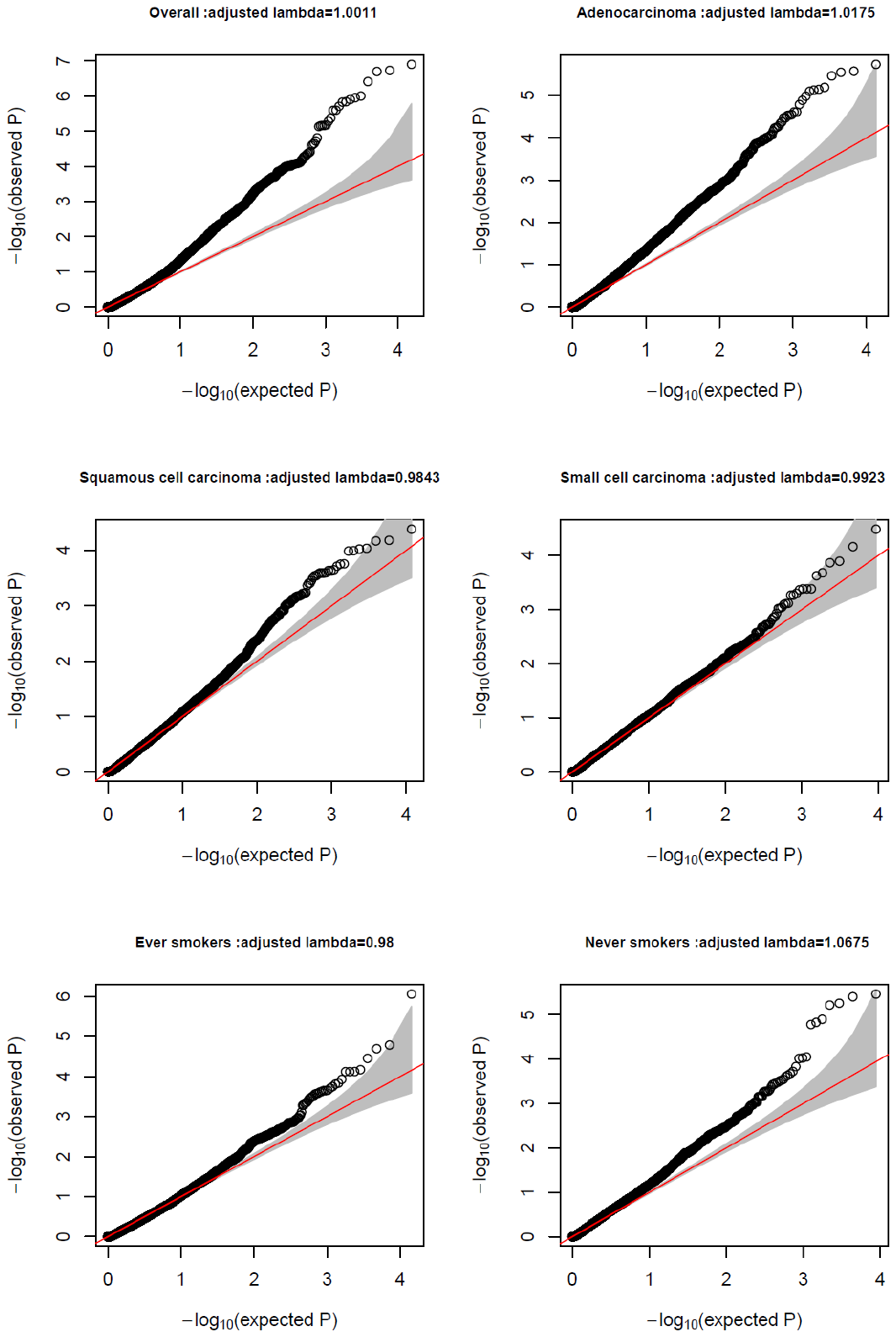

**Supplementary Figure S4. Quantile-Quantile plots of P-values from overall lung cancer association analysis and stratification analyses by histological subtype and smoking status.**

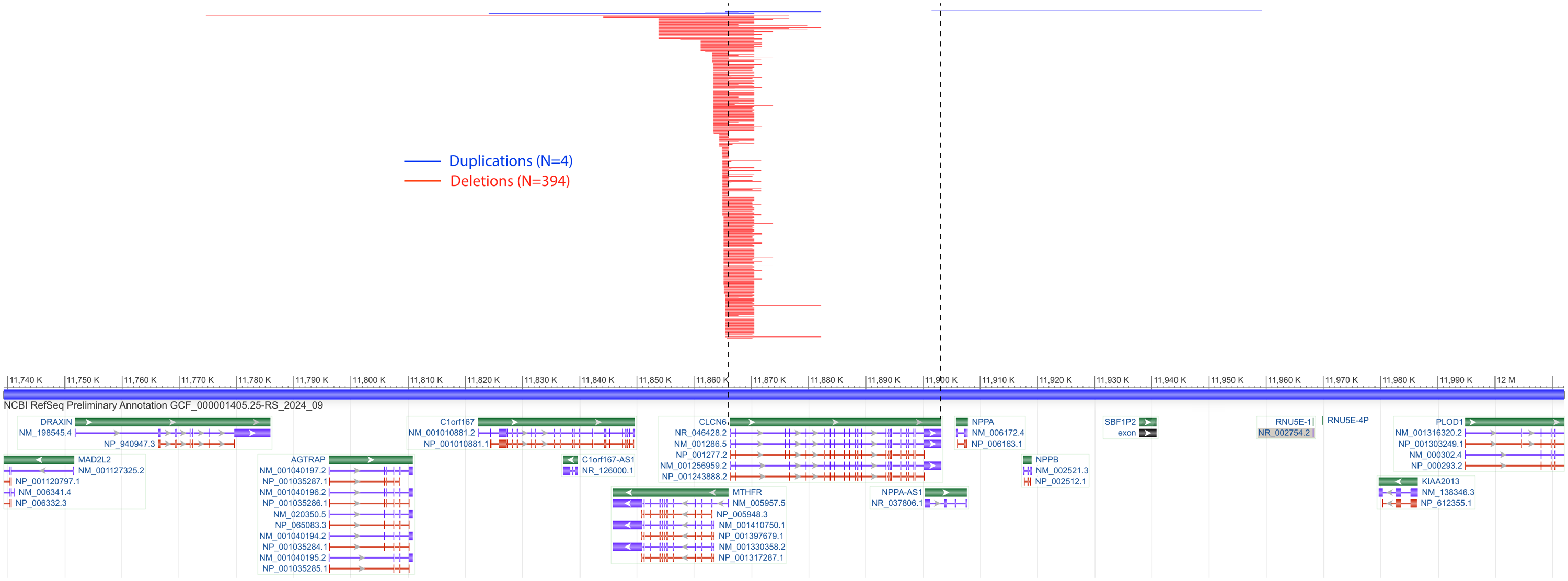

**Supplementary Figure S5. The physical positions of the identified copy number variants from the TRICL OncoArray samples in the 1p36.22 region.** 394 CNVs were identified in total including 4 duplications and 394 deletions, all overlapping with the gene coding region of *CLCN6* gene.

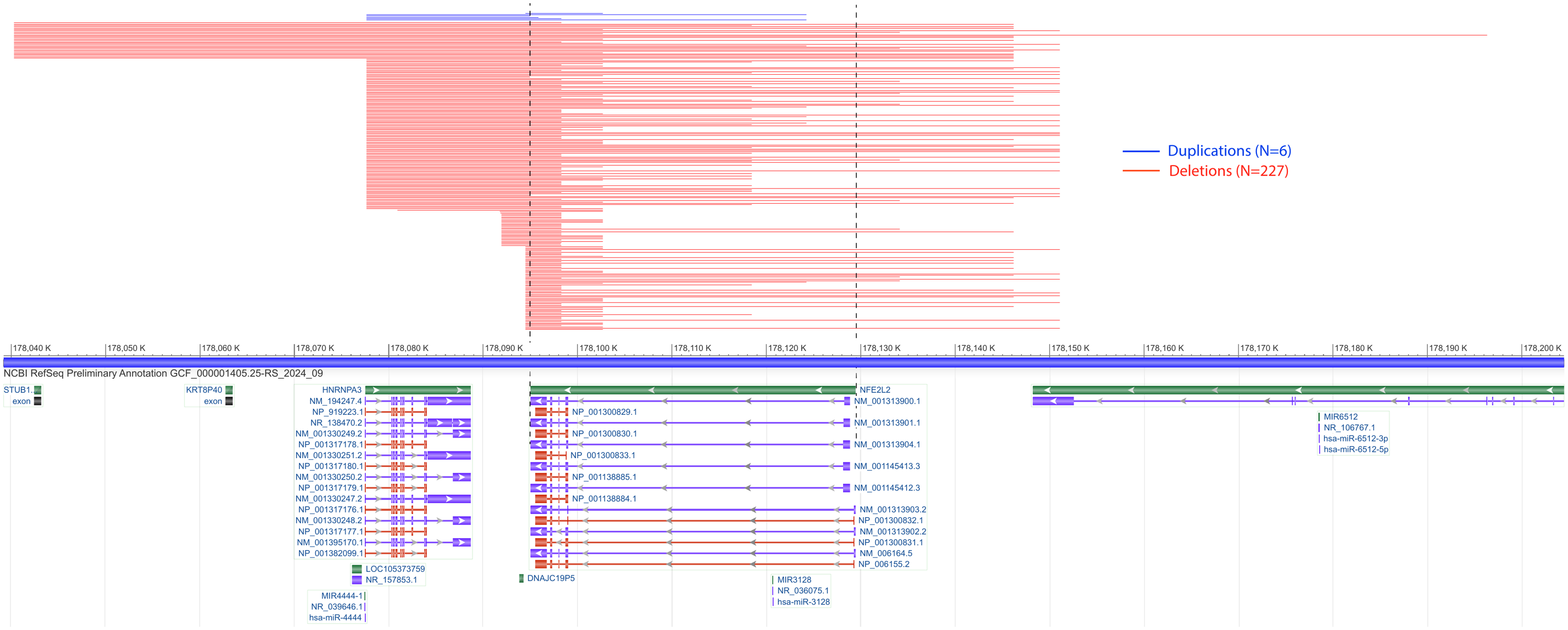

**Supplementary Figure S6. The physical positions of the identified copy number variants from the TRICL OncoArray samples in the 2q31.2 region.** 233 CNVs were identified in total including 6 duplications and 227 deletions, all overlapping with the gene coding region of *NFE2L2* gene.

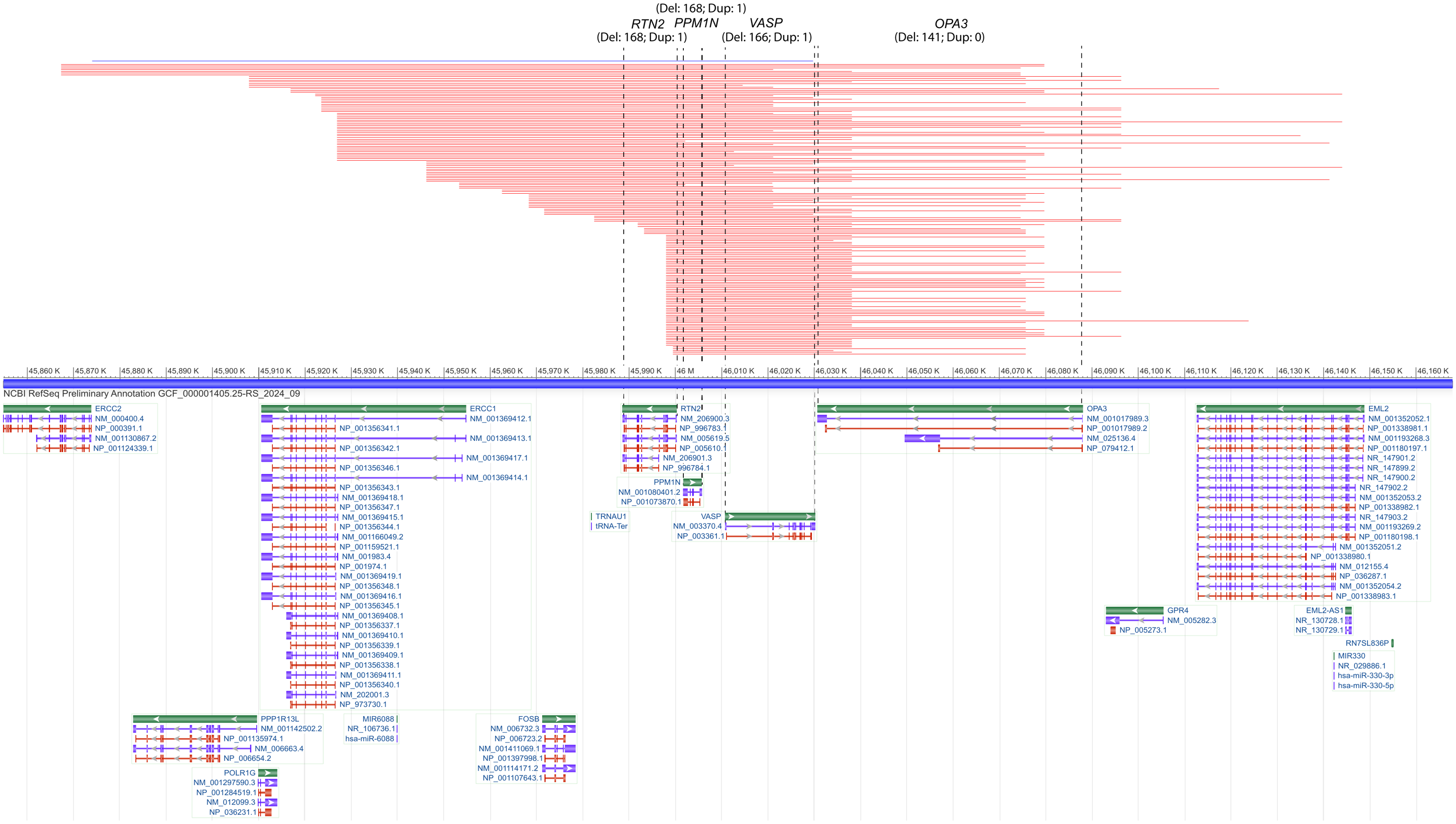

**Supplementary Figure S7. The physical positions of the identified copy number variants from the TRICL OncoArray samples in the 19q13.32 region.** These identified CNVs (total number of 168) are overlapping with the gene coding region of *RTN2*, *PPM1N*, *VASP* or *OPA3* genes*.*

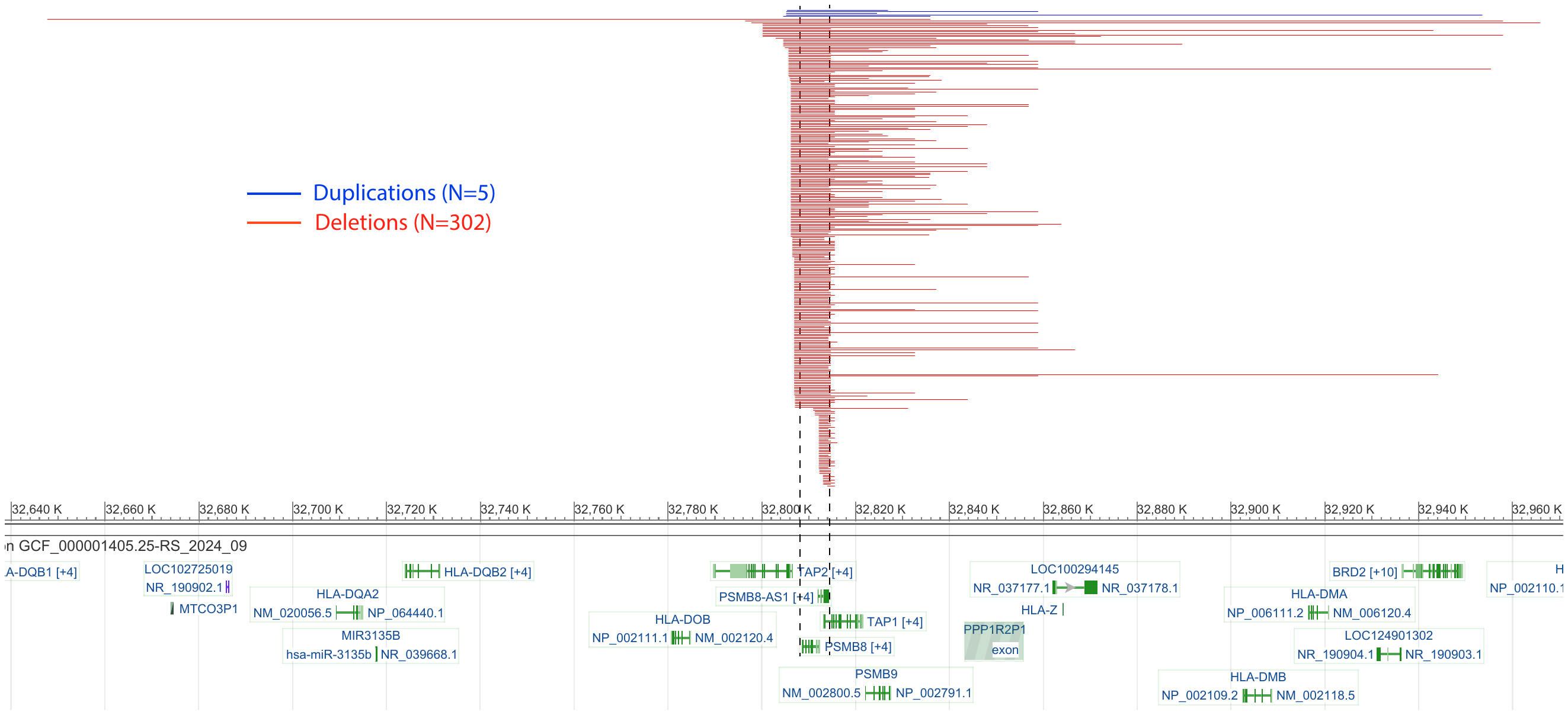

**Supplementary Figure S8. The physical positions of the identified copy number variants from the TRICL OncoArray samples in the 6p21.32 region.** 307 CNVs were identified in total including 5 duplications and 302 deletions, all overlapping with the gene coding region of *PSMB8* gene.

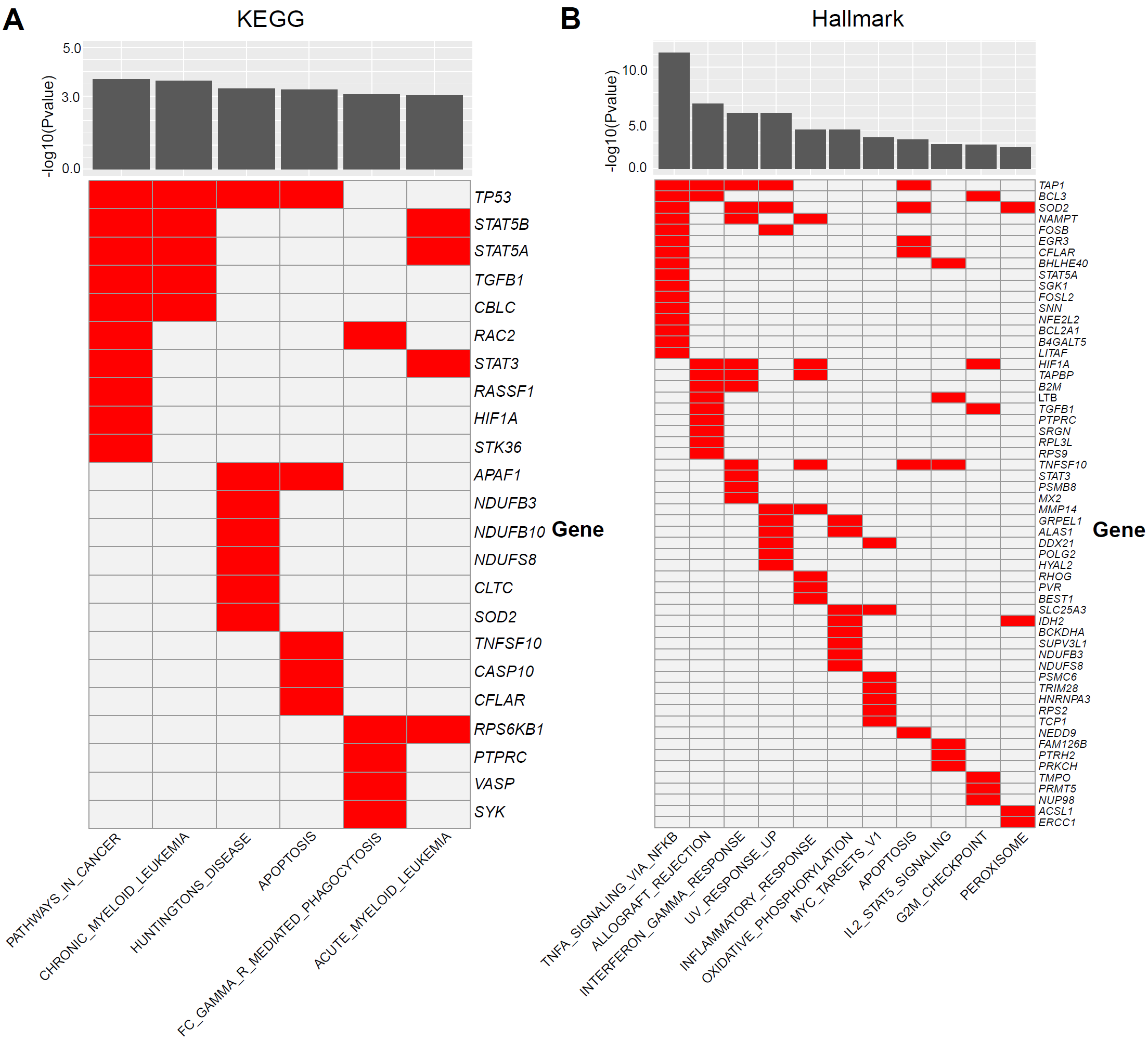

**Supplementary Figure S9. Pathway analysis based on significant gene sets.** Gene set enrichment analysis (GSEA) was performed on the top 200 CNV-associated genes in the gene-based and CNVR-based association analyses. Enrichment was tested against KEGG and MsigDB hallmark pathways. A false discovery rate (FDR) threshold of < 0.05 was used as a threshold for evaluating statistical significance.
